# Methods for Reproducible Comparison of Strategies in Stochastic Modelling

**DOI:** 10.1101/2025.10.09.25337145

**Authors:** Rob Sunnucks, Emma L Davis, Kat S Rock

## Abstract

Stochastic simulations are often used to model real-world phenomena such as infectious disease dynamics. In this modelling, differing strategies are often compared to one another by comparing the model outputs of each strategy. Hash-based matching, pseudo-random number generation is an approach for stochastic simulations that was implemented by Pearson and Abbott in the hashprng package to overcome challenges with comparing model simulations in a way that considers the dependency between model outputs. We demonstrate how methods based on this approach grant considerable benefit when comparing different strategies, and show when each of our three proposed methods ought to be used. We illustrate our methods with two epidemiological models: one simple model of a vaccine-preventable infection and one complex model of African sleeping sickness, which can be controlled through multiple interventions. We show how our Bernoulli hashing method works very well for simple models, and a variation of it can be used for more complex models in certain cases. Additionally, we discuss the properties of our methods for considering counterfactual scenarios and note that, compared to other attempts to obtain perfect counterfactuals, they demonstrate advantages in computational efficiency and their application to a wider variety of models.

## 1 Introduction

Stochastic simulations are often preferable to deterministic simulations in forecasting because they capture additional uncertainty that we expect in reality, beyond just parameter or cost uncertainty, and can produce integer outputs that are much more useful when considering discrete populations and phenomena such as elimination and extinction of infectious diseases.

When running stochastic models, the generated results are intentionally random, resulting in obtaining samples from a possible distribution of results. However, when attempting to analyse the effects of a specific change in the model, stochastic models have an issue in that they struggle to determine the statistical dependence between resulting distributions.

We are particularly motivated by examples of modelling the spread of an infectious pathogen through the use of a compartmental model. Specifically, we want to consider being able to compare different strategies to reduce infection transmission, which can be modelled by changing specific parameters in the system (e.g. introducing a vaccination rate, or reducing the force of infection— the rate at which individuals become infected).

### 1.1 Selecting the “best” strategy and health economics

We define a comparative metric as the difference or ratio between measures of two different, but comparable, systems. For example, we can measure the number of total cases under interventions A and B, and then have the number of cases averted by switching from intervention A to intervention B as a comparative metric. We note that in this paper, we will be using comparative metrics that are the difference of cumulative measures. If temporal measures are instead used, then outcomes may be ‘better’ or ‘worse’ at different times. For example, for “flatten the curve” interventions which reduce infectious contacts, we might expect there to be relatively fewer infections compared to no intervention during the middle of the outbreak, however relatively more infections at the end of the outbreak and a longer epidemic duration.

Other than the number of cases averted, other comparative metrics include the number of disability adjusted life years (DALYs) averted or net monetary benefit (NMB). A DALY is a weighted measure that combines the years of life lived with disability and the years of life lost due to disease. The NMB is a metric that combines the disease burden averted by a strategy with the additional cost of implementing it, by assigning a willingness to pay (WTP) value to avert a DALY.

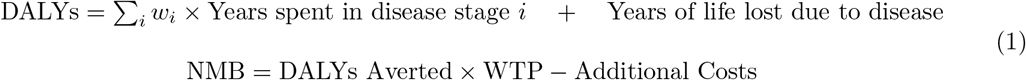

The disability weights *w*_*i*_ are dependent on the disease that is being modelled, with 0 representing a state of full health (i.e. no disability weighting for living with the disease), and 1 being equivalent to death. More information on the standardised weightings for different diseases can be found in the Global Burden of Disease Study [7].

Even with a well-defined metric, there are different methods for determining which strategy is better than the other. One of these methods is to calculate the expected value of undertaking each strategy. This is a risk-neutral approach, and regular stochastic simulations work fine asymptotically for this, due to the linearity of the expectation operator.

Another method, however, is to consider the probability that one strategy will be better than another. This is a more risk-averse approach. It is for methods like this that we consider more than just the expectation, where regular stochastic simulations will give noisy results. It is also worth noting that the standard error of the sample mean is the standard deviation divided by the sample size, and so regular stochastic simulations that give noisy results will also increase this standard error or may require more samples to obtain more accurate mean values.

Let us assume that we have an epidemiological model with *K* parameters, and we have *Q* parameter sets of these *K* parameters. This means our set of parameter sets is:

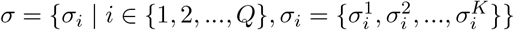

Let us assume we have two strategies, Strategy *A* and Strategy *B*. In order to determine the differences in effect for these two strategies, we will need some metric (e.g. the number of infections or NMB). We define 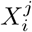 as the metric under parameters *σ*_*i*_ and Strategy *j* ∈ { *A, B* .}

Assuming each parameter set is equally likely, we then define the probability of Strategy *A* being optimal as:

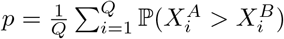

If we are using a deterministic model, then 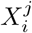 are deterministic variables. Therefore, *p* can be simplified as:

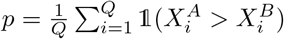

This works very well for the deterministic model, as 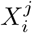 is only affected by parameter choices, which are matched for the compared simulations, and the strategy—which is what we are varying. For stochastic modelling, there is a third factor that affects the output—which is the random number generation during the simulation. If we are using a stochastic model, then 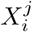 are random variables. When doing simulations, we get noise affecting our measurement for this value *p*.

For example, let us consider the simple example where, no matter what, Strategy *A* should always perform some *ϵ >* 0 better than Strategy *B*. i.e. 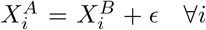. This means that we should have *p* = 1.

To determine *p* using simulations, we would run simulations to obtain *E* pairs of samples of 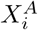 and 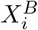 for each parameter set. These are 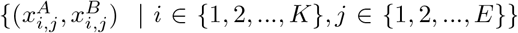 From these samples, we can calculate *p* as:

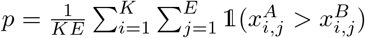

If we use a deterministic model, we will indeed find that 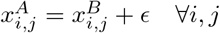 and so *p* = 1.If we run a typical stochastic model, then we can say that:

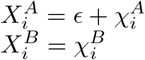

Where *ϵ >* 0, and 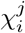 are random variables.

Because each compared sample is supposed to “take place in the same reality”, not only do we expect them to have the same parameter values, but we also expect them to have the same ‘underlying randomness’, i.e. 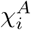 and 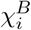 are dependent variables. However, in running our stochastic simulations, this dependence relation is not taken into account, and so these are effectively independent in our simulations, causing issues for our comparison.

Let us consider the case where *ϵ* = 0.5 and 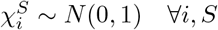. Then, in our regular stochastic simulations, where our metrics will be independent, we obtain that 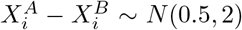, and so, 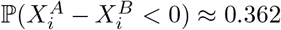 This means that instead of obtaining the correct result of *p* = 1, we find *p* ≈ 0.638. The independence of our paired stochastic realisations could incorrectly influence the decision-making process of the policymaker—although we believe Strategy A is always better than Strategy B and is parameterised in that way, our stochastic results indicate that it is not always better.

### 1.2 Counterfactual scenarios

These issues with stochastic modelling could result in a failure to fully estimate the impact of alternative interventions or situations. This could be in a retrospective manner, e.g. looking at what would have happened in the past if a different decision was made or if the situation was different. Alternatively, it could be in a prospective manner, e.g. how much better we expect one strategy to perform over another going forward.

The impact is therefore considerable. When we are looking at the success of previous interventions or the impact of past events beyond our control, it is vital that these are accurately captured in this assessment. We would not want to give evidence that stated there was considerable uncertainty as to the success of an intervention, when in fact the intervention was very likely successful. Additionally, if a new, future strategy is presented as making things worse with a high probability, when in fact the probability of this is much lower, policymakers may be less likely to adopt that beneficial strategy.

Outside of an infectious disease model example, we can generalise. Anytime we wish to obtain a comparitive metric using the resulting metrics of 2 similar models with a stochastic method, and care about more than the expectation of this comparitive metric, a regular stochastic simulation will give a noisy result. This demonstrates a need to find approaches that consider the dependence between compared samples, which is what we shall attempt to do with our reproducible stochastic modelling approach for counterfactual scenarios.

### 1.3 Existing approaches

Previous work has been done to obtain “perfect counterfactuals”; however, in trying to obtain perfect results, they are often unfeasible or unrealistic to implement into existing models, due to many changes needing to be made to the underlying model and large computation times. Work done by Kaminsky et al. on perfect counterfactuals for epidemic simulations [13] produced excellent results; however, this method is very computationally expensive and requires tracking each individual within the population. Work done by Cook et al. in constructing the effect of alternative intervention strategies on historic epidemics [4] requires the use of the Sellke construction, which uses individual thresholds of infection rather than standard compartmental modelling, and so is again not desirable for simulation of many epidemic models. Additionally, this method is better suited to models with time-homogeneous rates. Kaminsky et al. [13] pointed out similar issues in other articles, such as Kenah and Miller [15] not explicitly modelling timestructure and only having infection events, and Haydon et al. [10] not permitting counterfactual events that did not occur in the factual case. These present both issues when considering both future projections and retrospective questions.

We shall consider the work done by Abbott and Pearson in the development of Hashprng [22]. This is a package in R developed as a proof of concept of hash-based pseudo-random number generation as a compromise between perfect counterfactuals and computational run time. This involves hashing the current event and a salt value (which can be thought of as a base random seed that is shared between compared samples) and then using this hash to set the random number generator’s seed. This means that when events are the ‘same’ in compared samples, they will generate the same hash, and so undergo the same random process. This method is designed as an R package and does not yet have extensive amounts written about it. However, the underlying concept can be applied with ease to most models. Therefore, we will attempt to base our approach on the ideas used in the hashprng package.

We shall introduce three new approaches: the **default hashed model**, the **Bernoulli hashed model**, and the **truncated Bernoulli hashed model**. We will assess how well the default hashed model, the Bernoulli hashed model and the truncated Bernoulli hashed model perform at simulating comparisons of different strategies when compared to the standard stochastic model and the seeded model (2 existing, standard methods). We will illustrate the different approaches with two different epidemiological models: one simple model of a vaccine-preventable infection and one more complex model of African sleeping sickness, a vector-borne infection whose transmission can be controlled by medical and insect interventions. We will show how the Bernoulli method works very well for simple models, and a variation of it can be used for more complex models in certain cases. We also show that the hashed approach performs much better than simply seeding a stochastic model.

### 2 Methods

### 2.1 Overview and assumptions

The tau-leaping method was devised in 2001 [8] as an alternative approach to the Gillespie algorithm for simulating mechanistic models with stochastic changes to a discrete collection of individuals. Tau leaping provided sizeable reductions in computation time compared to the Gillespie algorithm, with a tradeoff of a slight reduction in accuracy. Since then, tau-leaping has been used in many fields, from kinetic chemistry [8] to ecological and infectious disease modelling [16]. To summarise, the tau-leaping algorithm works by stepping through time in discrete time steps. After each time step, the number of each of the events that occured in that time step are drawn from a Poisson distribution, with parameter equal to the rate of the event multiplied by the length of the time step. Then the events occur, updating the population. Tau-leaping can lead to issues when outflows from one source split into different destinations. Should this be the case, and enough events occur such that the source population falls below 0, it must be decided which events occur, and which do not. Typically, this is decided using a multivariate hypergeometric distribution (note that for our methods, these must also be seeded using the hashing function). The **regular stochastic simulation** we shall consider involves performing this tau-leaping process, where the number of each event is drawn randomly from a Poisson distribution every time step. The most common step taken to try to implement counterfactual simulations is the **seeded stochastic method**, where a random seed is set once before each simulation, such that compared realisations had the same initial random seed.

We are going to introduce alternative methods which use hash functions to set the random seed throughout the simulation, based on work done in hashprng[22]. The most simple of these we refer to as the “**default hashing method**”, which sets the seed to the result of a hashing function before each Poisson event draw. A hashing function maps a set of data into a single string of bits called the hash value. Collisions occur when a hash function maps two different sets of data to the same binary string. A good hashing algorithm will minimise the probability of collision. For the work in this paper, the hash value is used to set the random seed of the system before each event draw. By having fewer collisions, we reduce the probability of reusing the same random seed. We then consider the **Bernoulli hashing method**, which replaces the Poisson event draw from the default hashing method with a sum of Bernoulli random variables, in an attempt to make a ‘pseudo-individual’ model. Additionally, we shall look at the **truncated Bernoulli hashing method**, which pulls events from the tail end of the Bernoulli distribution after the hashing sets the random seed to reduce computational complexity. We note that another method, the state-based hash, was also considered but is not presented in the main text as it is not deemed as useful as the other methods for the epidemiological models considered here. Information on this can be found in Section 2 of the Supplementary Information. Further information on the coding of these methods can be found in Section 3 of the Supplementary Information.

A summary of the various methods can be seen in Table 1.

**Table 1:** Comparison of the various methods.

|  | **Default Hashing** | **Hashed Bernoulli** | **Truncated Bernoulli** |
| --- | --- | --- | --- |
| Distribution of number of events | Poisson | $\sum$ Bernoulli | $\sum$ Bernoulli |
| Dependence on the number of events drawn between realisations | Always dependent and ‘proportional’ | Dependent between realisations but not ‘proportional’. Individual Bernoullis are ‘proportional’. | Dependent between realisations, but underlying beta and uniform distributions are ‘proportional’. |
| Events can be resolved inconsistently | No | No | Yes, with very low probability, which tends to 0 as the time step tends to 0. |
| Computational speed | Quick | Slow (scales with population size) | Quick (scales with truncation point) |

Let *X*_1_, *X*_2_ be a pair of random variables, with cumulative distribution functions *f*_1_(*x*) and *f*_2_(*x*) respectively. We say *X*_1_ and *X*_2_ are proportional iff

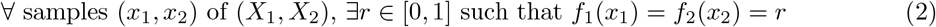

We consider an event to be a number of well-mixed, interchangeable individuals in a compartment, experiencing certain outcomes resulting in them changing compartments in a time step. Additionally, we assume the events that occur at time step *t* depend solely on the state of the system at time step *t*. Therefore, the only dependence between events is that previous events can alter the future state, and the state alters the rate at which events occur. However, given a rate, the underlying random process used to generate the number of events that occur at a specific time step is not changed by the events at other time steps. We state that the consistent resolution of events requires that, for a given realisation (salt value), at a given time step, the number of events that occur must not decrease with the rate of that event. To formulate this mathematically:

Let *E*_*i,t,s*_(*λ, n*) be the number of times event *i* occurs during time step *t*, for a realisation using salt value *s*, where the rate of an individual undergoing an event is *λ*, and *n* individuals can undergo the event. Then,

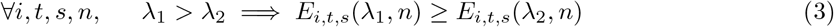

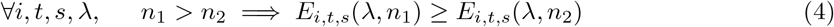

We have assumed that the deterministic, continuous system of ODEs that uses the same rates as our stochastic system is not chaotic, which is true for the majority of compartmental infection models. Here we are defining a system to be chaotic if it has sensitive dependence on initial conditions, i.e. given the trajectory from any set of initial conditions, the trajectory of a set of nearby initial conditions will diverge exponentially. Examples of chaotic systems include the Lorenz system used to describe weather [17, 6]. For example, in systems we consider, an individual getting vaccinated does not lead to drastically different behaviour in the rest of the population. If this system were chaotic, then we would expect issues to arise, as our assumptions may not be realistic.

We note that this means that in the long term, it is possible (however quite unlikely) for events that should be beneficial on an individual level basis to result in cumulatively worse outcomes. For example, consider a simple Susceptible-Infected-Recovered (SIR) model, and suppose we start out with one initially infective individual. We consider an improved strategy, where the rate of transmission is slightly reduced compared to a baseline strategy. Under the improved strategy, the first five time steps may have no infections and no recoveries each time step, whilst under the baseline strategy, there may have been one infection each time step, and one recovery in every time step except the first. Suppose then in time step six, we have no events under the improved strategy, but we have one recovery event under the baseline strategy. This means, after six time steps, we have one infected and no recovered under the improved strategy, and one infected and five recovered under the baseline strategy. In the next time step, we could draw a recovery event for both strategies, but only an infection event under the improved strategy (as there are more susceptibles). This would mean the outbreak has concluded under the baseline strategy (infecting six people in total), but can continue under an improved strategy, where it may go on to infect more than six individuals. This is effectively a delayed take-off, which could result in more infections due to the randomness of the future being different from the randomness of the past. Under both strategies, that same future randomness exists, but it only affects the outcome of the improved strategy, as only the improved strategy still has infections remaining at that point. We consider this to be a realistic possibility for our model to include, albeit an unlikely one.

## 2.2 Default Hashing

The default hashing algorithm is based directly on ideas used in the hashprng package by Peason and Abbot [22]. Under this approach, the tau-leaping process still draws Poisson random numbers to determine the number of events of each type. However, before each of the random number draws, we shall set the random seed to be the result of a hashing function. This ensures that counterfactual realisations have the same random seed at each time step, resulting in a statistical dependence between these two realisations.

The hashing function takes in the time step, salt value, and the event type to produce a hash value. The salt value should be the same between counterfactual realisations, but unique otherwise. For example, if we wish to simulate *C* trajectories, we define a set of unique salt values 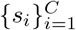, and set the salt value of the *i*^th^ simulation of each counterfactual scenario to be *s*_*i*_. This then means that the *i*^th^ trajectory in each scenario can be compared.

As an example of what this might look like, suppose we have two compared strategies—A and B, and the rate of recovery is the same for both. For a realisation of Strategy A, we have 10 infected individuals at time *t*, and for B, we have 20. If in A’s realisation, we obtain two recoveries, we should obtain (approximately) four recoveries in B’s realisation. Additionally, if we had the same number of infected individuals under strategies A and B, then we would obtain the same number of recoveries in both of their realisations.

We should also note that time has to be passed into the function. Let us consider reaching a point in time, *T*, where no events are taken in the next time step. This means, if time is not included in the hashing function, the hash values will remain the same at the next time step, and so again no events will take place. This would result in the simulation state becoming constant and not changing in time. The possibility of reaching a point in time with no events is also more likely with lower rates and time steps. Hence, time should be included in the hashing function to prevent this phenomenon.

We do note that there exists a ‘proportionality’ between realisations as a statistical property of the default hashing method. This is because each compared realisation is drawing from the same percentile of the Poisson distribution. The previous example of the recovery event illustrates this. In reality, this may not be the case.

We note that the behaviour of proportionality ensures that having more individuals or a higher individual event rate in a compartment does not lead to fewer events (and vice versa). Indeed, if we have *N* individuals in a compartment under Strategy A at time *t* and *N* + *n* in Strategy B at time *t*, then the number of events of individuals leaving this compartment will be between 0 and *n* greater in Strategy B than in Strategy A. This ensures the consistent resolution of events.

It should be noted that under specific, unlikely conditions, e.g. if the number of random numbers drawn at every time step is always the same, a default hash may be equivalent to setting the random seed beforehand.

### 2.3 Bernoulli Hashing

The next approach we consider is a Bernoulli-based hashing algorithm. This extends the default hashing method by keeping the hashing, but changing how the number of events is calculated in an attempt to remove this ‘proportionality’. The tau-leaping process no longer draws Poisson random numbers; instead, it draws a sum of Bernoulli random numbers. If there are *N* possible individuals that might undergo an event, it draws a Bernoulli random number to determine if each of them undergoes the event. In doing this, we allow for each individual in the compartment to behave similarly between compared realisations, as the uniform random number behind each Bernoulli draw will be the same.

The probability of an individual undertaking an event in a time step *τ* with individual rate *λ* is *p*(*λ, τ*). We base this on the Poisson construction, with one individual in a compartment. The probability they would undergo an event is 1 − ℙ (0 events occur), and so: *p* = 1 − exp(−*λτ*)

For example, suppose we have compared realisations for Strategies A and B, and the rate of recovery is the same in both. In the realisation for Strategy A, we have 10 infected individuals at time *t*, and in the realisation for Strategy B, we have 20. If we draw *x* recoveries in A’s realisation, then in B’s realisation we will draw *x*+*Y* recoveries, where *Y* ∼ *Binomial*(10, *p*)—unlike the default hashing where *Y* is proportional to the extra individuals under Strategy B. If instead we had the same number of infected individuals in the realisations of both strategies, then we would obtain the same number of recoveries in both realisations.

It should be noted, the random seed is only set once per event, not once for each individual in the compartment times, and the hashing function used for the Bernoulli method behaves the same as in the default hashing method.

The Bernoulli method does not fully remove the dependence between realisations, but does remove ‘proportionality’, as only shared individuals have the same random behaviour. It also ensures the consistent resolution of events. Firstly, more events will not occur in a realisation with fewer people who can undergo that event. To see this, let us have two scenarios, A and B, with the same individual event rates, with *N*_*A*_ *> N*_*B*_ individuals in a compartment, and let *b*_*i*_ ∈ { 0, 1} be our Bernoulli random variables. Then:

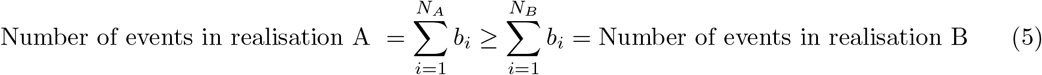

Secondly, more events will not occur in a realisation with a lower individual event rate. To see this, let us have two scenarios, A and B, with the same number of individuals in a compartment, *N*, with *p*_*A*_ *> p*_*B*_ as the probabilities of each individual undergoing the event. Let *u*_*i*_ ∈ [0, 1] be the uniform random numbers used to obtain our Bernoulli random numbers. Then:

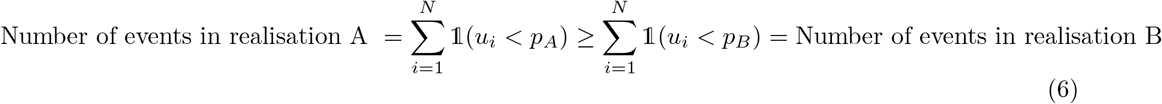

The downside of this method is that you must draw a Bernoulli random number for each possible event, which becomes very computationally expensive for large population sizes or very complex models with a lot of different events.

### 2.4 Truncated Bernoulli Hashing

The potentially large computation time of the Bernoulli method can present practical implementation issues. To remedy this, we present a variation of the Bernoulli method—the truncated Bernoulli method. This is designed to replicate the behaviour of the Bernoulli method for larger and more complex models that have low rates, by drawing a limited number of Bernoulli random numbers from the tail of the distribution.

We once again keep the same hashing method, setting the random seed to be the result of the hashing function. For the normal Bernoulli hashing method, we generate **X** = (*x*_1_, *x*_2_, *x*_3_, …, *x*_*N*_) ∈ {0, 1}^*N*^, where each *x*_*i*_ is a Bernoulli random variable with some probability *p*. In practise to do this, we actually generate **R** = (*r*_1_, *r*_2_, *r*_3_, …, *r*_*N*_) ∈ [0, 1]^*N*^, where each *r*_*i*_ is a uniform random variable between 0 and 1, and then set: *x*_*i*_ = 𝟙(*r*_*i*_ *< p*) then we take the number of events to be 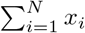 .

Consider the case where 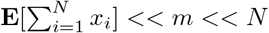, for some *m* ∈ **N**. Then we define an ordering *σ* such that *r*_*σ*(*i*+1)_ ≥ *r*_*σ*(*i*)_ ∀*i*. We can note that *x*_*σ*(*i*+1)_ ≥ *x*_*σ*(*i*)_ ∀*i*. Therefore that all the non-zero values of our Bernoulli *x*_*i*_ are (very likely to be) in 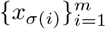 If we can find this 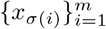, that will be sufficient to determine the number of events, 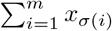 To do this, we note that the *r*_*σ*(*m*)_ is the *m*^th^ order statistic of the uniform distribution, which is a beta-distributed random variable:

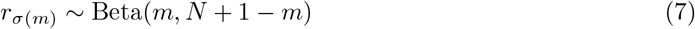

Now that we have calculated the largest of our uniform random numbers, we can generate the remaining ones with a simple uniform distribution:

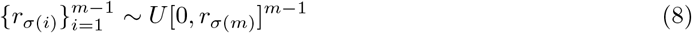

Finally, we can obtain the number of events that occur in this timestep:

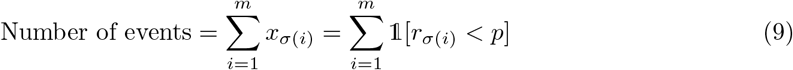

This truncated Bernoulli method will be substantially faster than the initial Bernoulli hashing method for complex models, having to only draw *m << N* random variables now. The truncated Bernoulli does lose some of the nicer statistical properties of the Bernoulli method, however.

Consider realisation *i* of Strategy A, where we have *N* (large) susceptibles, and realisation *i* of Strategy B, where we have *N* + *M*. Under the Bernoulli approach, if we have *k* infections under Strategy A, we must have between *k* and *k* + *M* infections under Strategy B. Under the default hashing approach, if we have *k* infections under Strategy A, we must have 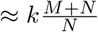 infections under strategy B. However, in the truncated Bernoulli approach, if we have *k* infections under Strategy A, we must have between *k* and *m* infections under Strategy B.

This leads to the possible example of *M* = 1, *k* = 0, that is, for Strategy A, we have *N* susceptibles, and no infection events and for Strategy *B*, we have *N* + 1 susceptibles. Under the Bernoulli and default hashing approaches, we can only have 0 or 1 infections for Strategy B, as these approaches have consistent resolution of events. However, with the truncated hashing approach, we could have between 0 and *m* infections. Therefore, in incredibly rare circumstances, we can reach our maximum number of infection events by adding an individual to the susceptible population. This demonstrates that this method does not always resolve events consistently; however, the probability that events are resolved inconsistently is low and decreases if the time step and/or individual event rate decreases. A more detailed explanation of how this can occur can be seen in Section 1 of the Supplementary Information.

For the truncated Bernoulli, it is also possible that an event is missed due to the truncation. Of course, this is more of a concern when the expected number of events in a given time step is close to *m*. The probability of this can be calculated, and we shall look at how it varies with the size of the time step taken, as the size of the time step is proportional to the expected number of events in a time step.

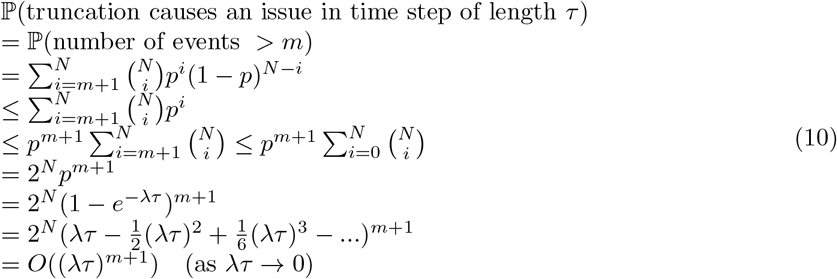

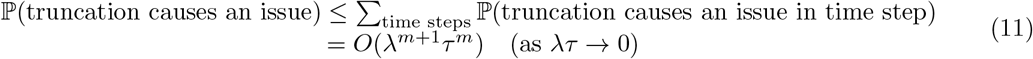

This probability tends to 0 as *τ* tends to 0, and decreases as we increase the value of *m*. However, it should be noted that increasing *m* will also increase the computation time.

How small/low the time interval *τ* and rate *λ* should be to use the truncated Bernoulli depends on the maximum number of events, *m*, we allow in a time step. The probability of an event occurring for each possible person is dependent on *λτ*, and the truncated model fails when the number of events drawn in a time step would exceed *m*. Hence, a binomial distribution can be used to determine the probability of model failure, as seen in the above calculations. Larger values of *m* allow for more events in a given time step (and so a lower probability of model failure), but will also take longer to simulate. This is the effective trade-off. The user of the method can determine what they believe is a reasonable probability of method failure, and determine their value of *m* accordingly.

## 3 Applications

### 3.1 Simple Example: SEIRV

We consider a simple Susceptible-Exposed-Infected-Recovered model [14], with an additional vaccinated class added on (SEIRV). The transitions in this model are exposure, becoming infectious, recovering and becoming vaccinated. A diagram of the model can be seen in Figure 1, which shows the progression of infection and how vaccination occurs. Further information on the model can be found in Section 4 of the SI.

**Figure 1:**
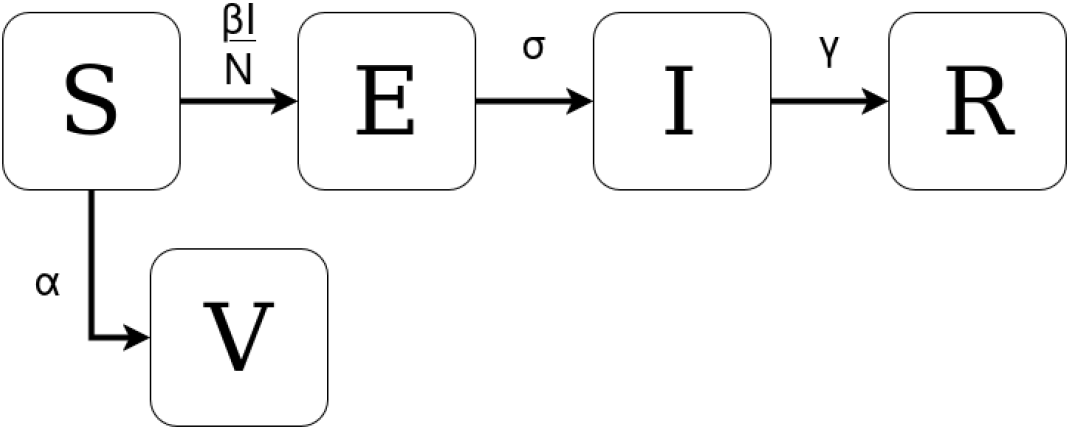
The compartmental SEIRV model with arrows showing infection progression and associated rates.

In our analysis we selected the following mean parameter values, *θ* = (*β, σ, γ*) = (1, 1, 0.45). The parameter *α* corresponds to the strength of the vaccination strategy and is varied accordingly. Using this model, we wish to determine the number of infections averted by implementing a vaccination intervention compared to continuing with no vaccination. We consider an outbreak infection, starting with 10 infected people in a population of 10,000 people, and with no demography. Without an intervention we would expect a large outbreak, with the mean peak prevalence of approximately 25%, infecting an average of 68% of the population by the end of the outbreak.

To include parameter uncertainty, we will generate 10,000 sets of parameter values, *θ*_*i*_, and for each of these we can run a simulation to obtain the number of infections under a vaccination strategy 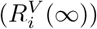, and the number of infections under a no vaccination strategy 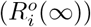 across a 1000 day time horizon. Then, we calculate the infections averted as:

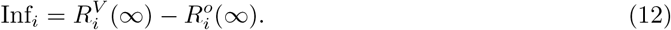

We can compare how the distribution of Inf_*i*_ changes as we vary both the vaccination rate and the simulation method; we will conduct simulations using the ODE formulation and the different variants of the tau-leap stochastic simulations [24]. The variance of Inf_*i*_ will be used to show the uncertainty in the modelling method. This model was made using MATLAB version R2024a [12], and random number generation was done using the default random number generating functions.

In Table 2 we can see the average time taken per simulation for each of the methods. It should be noted that this is variable. We do note that the new methods seem to be getting faster (relative to the regular stochastic method) as we increase the size of the time step. Firstly, Figure 2, show the infections averted for our different simulations where a time step of 0.25 was used. Figure 3 shows the variance for each of these strategies to allow us to quantify the uncertainty for different simulation types.

**Table 2:**
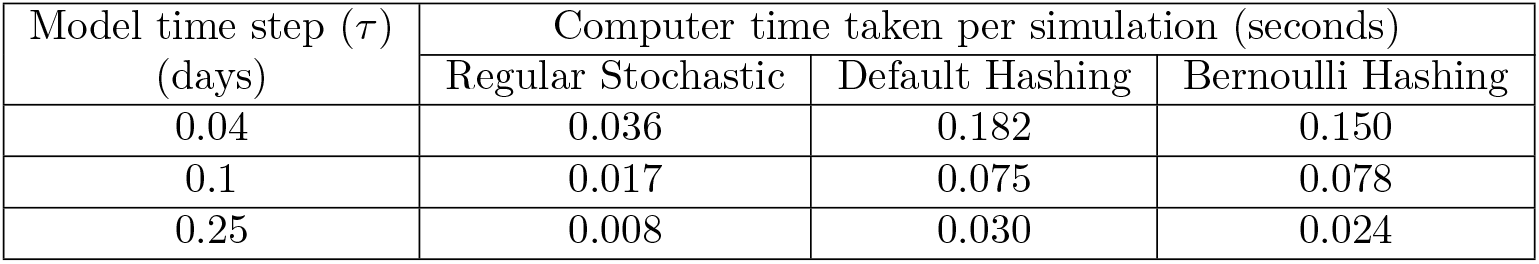
Comparison of the computer time taken per simulation of the SEIRV model under three different methods and three different model time steps. Computer time is in seconds and is given to 3 decimal places.

| Model time step ($\tau$)<br>(days) | Computer time taken per simulation (seconds) | | |
| --- | --- | --- | --- |
|  | Regular Stochastic | Default Hashing | Bernoulli Hashing |
| 0.04 | 0.036 | 0.182 | 0.150 |
| 0.1 | 0.017 | 0.075 | 0.078 |
| 0.25 | 0.008 | 0.030 | 0.024 |

**Figure 2:**
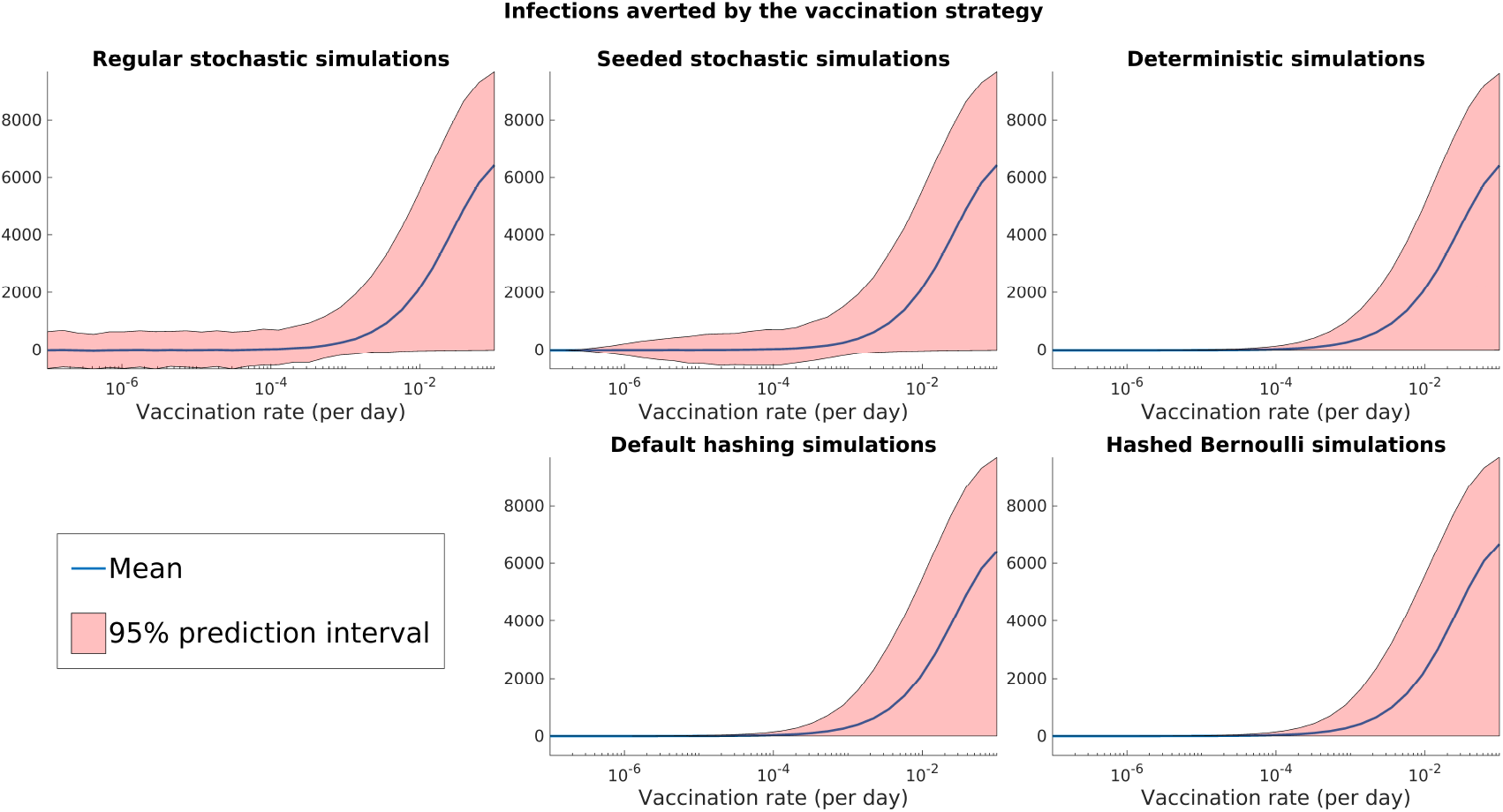
Plots of the infections averted by undertaking vaccination, where vaccination rate is varied, for our five different simulation methods. Stochastic simulations used a value of *τ* = 0.25 for these results. A quantification of the variance of these results can be seen in Figure 3.

**Figure 3:**
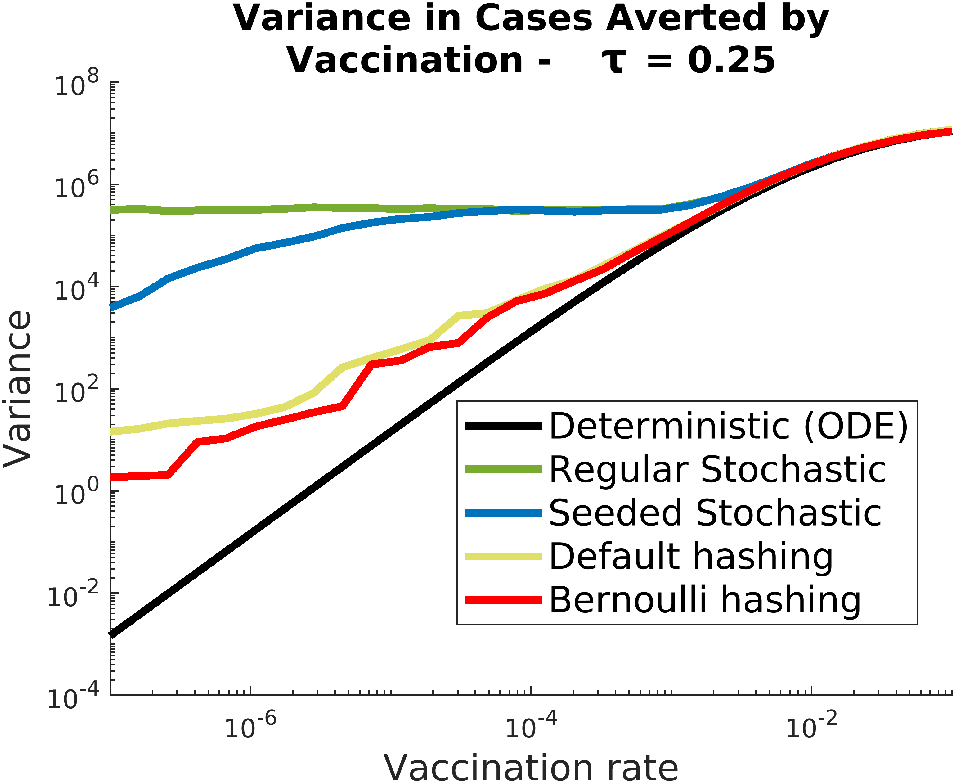
Comparison of the variance of infections averted for various simulation methods, with a value of *τ* = 0.25.

**Figure 4:**
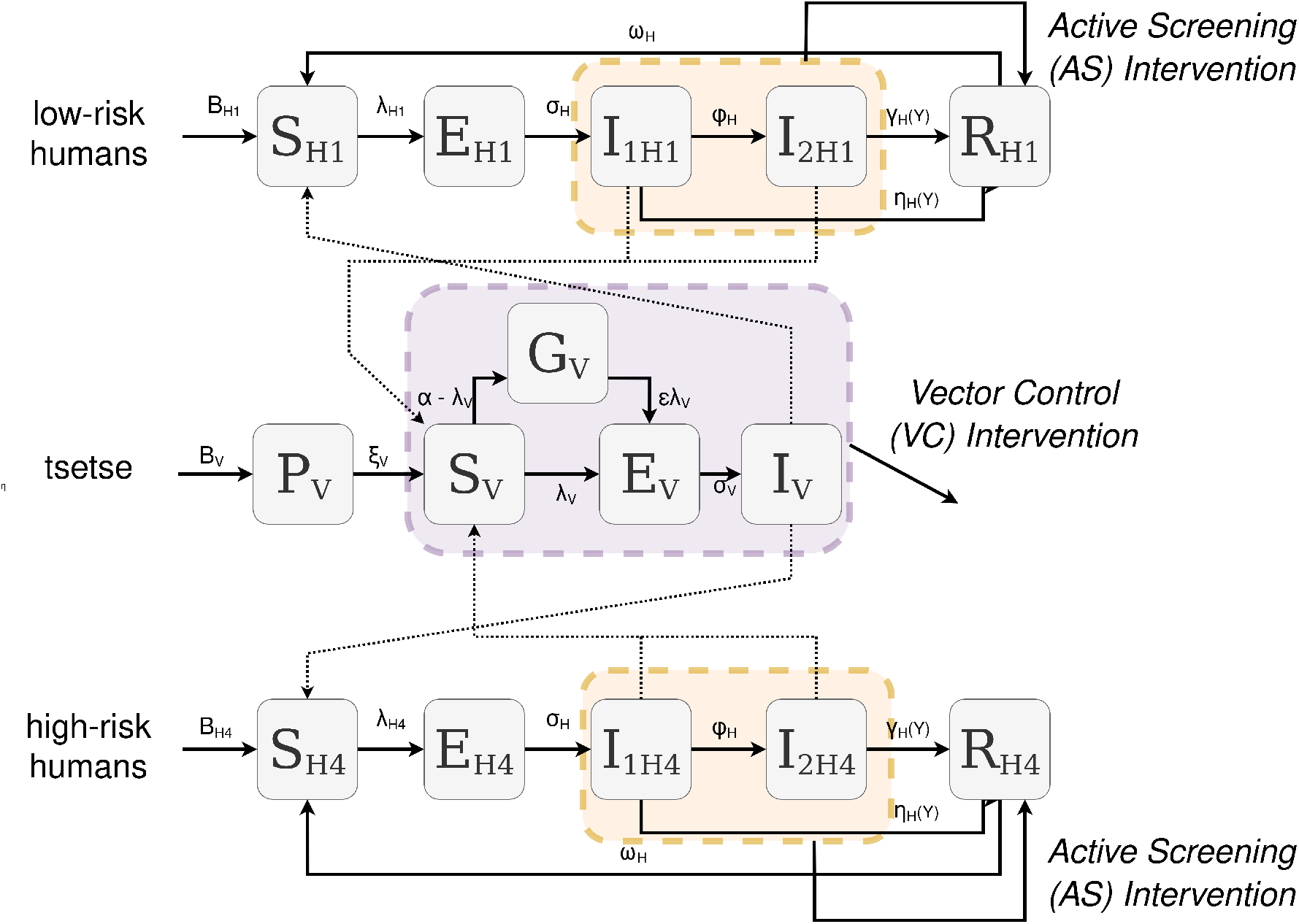
Illustration of the gHAT compartmental model (adapted from Antillon et al. [2] under a CC-BY license).

We can see that the use of hashing in our stochastic simulations has reduced the uncertainty on the infections averted by vaccination. This is more noticeable for lower vaccination rates, as higher vaccination rates lead to much more noise from other sources, such as parameter uncertainty, and so the stochastic noise is smaller in comparison.

Overall, we can see that the two hashing methods presented provide large improvements to the regular stochastic and seeded stochastic models. The regular stochastic method appears to have a very high level of uncertainty that does not decrease as the vaccination rate decreases (i.e. as the strategies become more alike). The seeded stochastic method provides a small improvement on this, with it starting to decrease once the vaccination rate becomes very small. The default hashing method then provides a large reduction in the variance at the cost of taking a few times longer to run (exact slowdown depends on the size of the time step *τ*). Finally, for a model as simple as this, the Bernoulli method gave the largest reduction in variance—being slightly better than the default hashing method, whilst still running in a reasonable time, taking about as long as the default hashing method.

Additionally, both of these hashing methods seem to, at least when looking at their 95% prediction interval, not result in negative infections averted due to vaccination. This differs from the seeded and regular stochastic approaches, which do get these negative values, which would imply that vaccinating the population could cause more infections—which is not reasonable.

In order to compare these different approaches to a “ground truth” we coded the Kaminsky et al. method for our SEIRV model. Whilst this code does run, we found that it is prohibitively slow to run the same set of tests to obtain comparable results to the other proposed methods. The RAM requirements would be around 2 TB, and it would take a several years to run. Even if efficiencies are made in the code, it is impractical for us to make such a comparison in the present study. Full code for our implementation of the Kaminsky method for the SEIRV model alongside all other code used in this study can be found at Open Science Framework (https://osf.io/u74h3/overview).

### 3.2 Complex Example: gHAT

*Gambiense* human African trypanosomiasis (gHAT), is on the World Health Organization’s (WHO) list of neglected tropical diseases (NTDs) [20], is targeted for elimination [18], and is endemic in 24 countries of West and Central Africa[19]. gHAT is a vector-borne disease, caused by a protozoan parasite spread by the tsetse, a biting fly which has unique biology [29]. The gHAT model was developed by the HAT Modelling and Economic Predictions for Policy (HAT MEPP) research project [5] [11] and is much more complicated than the SEIRV model above. However, the endemic infection prevalence in the model is very low; a 1% prevalence would even historically be considered extremely high prevalence for gHAT [26]; in this example, there is around a 3.2% mean prevalence at the beginning of the simulation (the year 2000), or around a 0.08% mean prevalence by the time we consider diverging strategies (the year 2025). In this illustrative example, we assume a population size of 180,000 people.

To simulate different strategies to compare, we will consider two possible levels of active screening (AS): mean (low) and max (high), and two levels of vector control (VC): targeted (low) and full (high). AS [23] involves sending mobile teams to the region to test people for gHAT on a large scale, with those who are confirmed as having the infection then receiving treatment. The mean level of AS assumed here involves screening 33% of the population each year, whilst the max level involves screening 50% of the population. VC is the deployment of Tiny Targets, which are devices that tsetse are attracted to that have been impregnated with insecticide, leading to a sizeable increase in mortality for tsetse, reducing their population [28]. This then results in four total strategies, which are combinations of these two interventions. In all scenarios there is continued surveillance in fixed health facilties (known as passive screening) which relies on sick people presenting themselves to trained health workers who have available diagnostic tests. Based on real-world guidance [18], we assume that AS and VC would stop after three years with no reported cases, even if there are still remaining infections in the model. We will then investigate two different metrics for the success of the intervention. One of these is the net monetary benefit (NMB), measured in dollars and balances the cost of an intervention with the disease burden prevented (measured in DALYs). The cost function for calculating the NMB can be found in Section 5 of the Supplementary Information and uses known cost data [1][25]. The second metric of interest here for gHAT is the predicted year of the last transmission event (LTE); similar to the calculation of DALYs which needs the model to estimate deaths and disease burden, it is impossible to know from direct observation whether the LTE has already occurred but it can be estimated from model outputs [27].

For the simulations, we will run 5000 realisations for each strategy from the year 2000 until 2055, with the change in active screening levels and introduction of vector control being introduced in 2025. We selected this time horizon to capture the impact of the various strategies if they were to be implemented in the year 2025 and such that we could be sure to observe the LTE. The time period of 2000 to 2025 serves the purpose of getting us varying initial conditions for the future projections. This was simulated using C++ code run inside a MATLAB wrapper code, and GSL [9] random number generation was used. Full code for our implementation of the gHAT model alongside all other code used in this study can be found at Open Science Framework (https://osf.io/u74h3/overview).

We would expect that increasing the levels of AS and/or VC would result in fewer DALYs and an earlier LTE. This is because AS reduces the infected human population, and VC reduces the tsetse population (including infected tsetse), both of which therefore reduce the force of infection. Indeed, this is what we have observed in our results—with increasing VC clearly leading to an earlier LTE, as seen in Figure 5, and both increased VC and increased AS leading to further DALYs averted, as seen in Figure 6. The seeded stochastic simulations often have increasing VC resulting in a later LTE. When using hashing methods, this does not occur, which is reflective of what we would see in reality.

**Figure 5:**
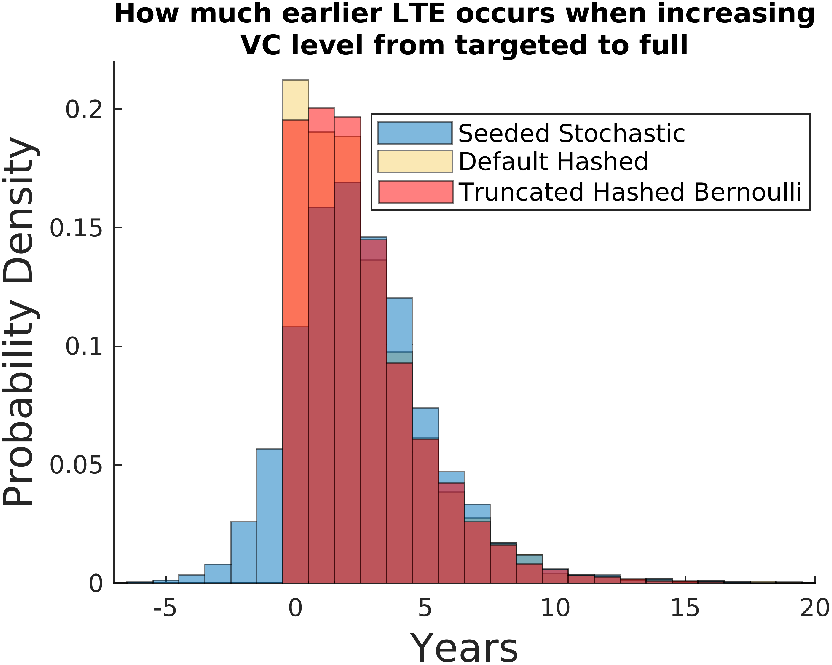
How much earlier LTE occurs by increasing the level of VC

**Figure 6:**
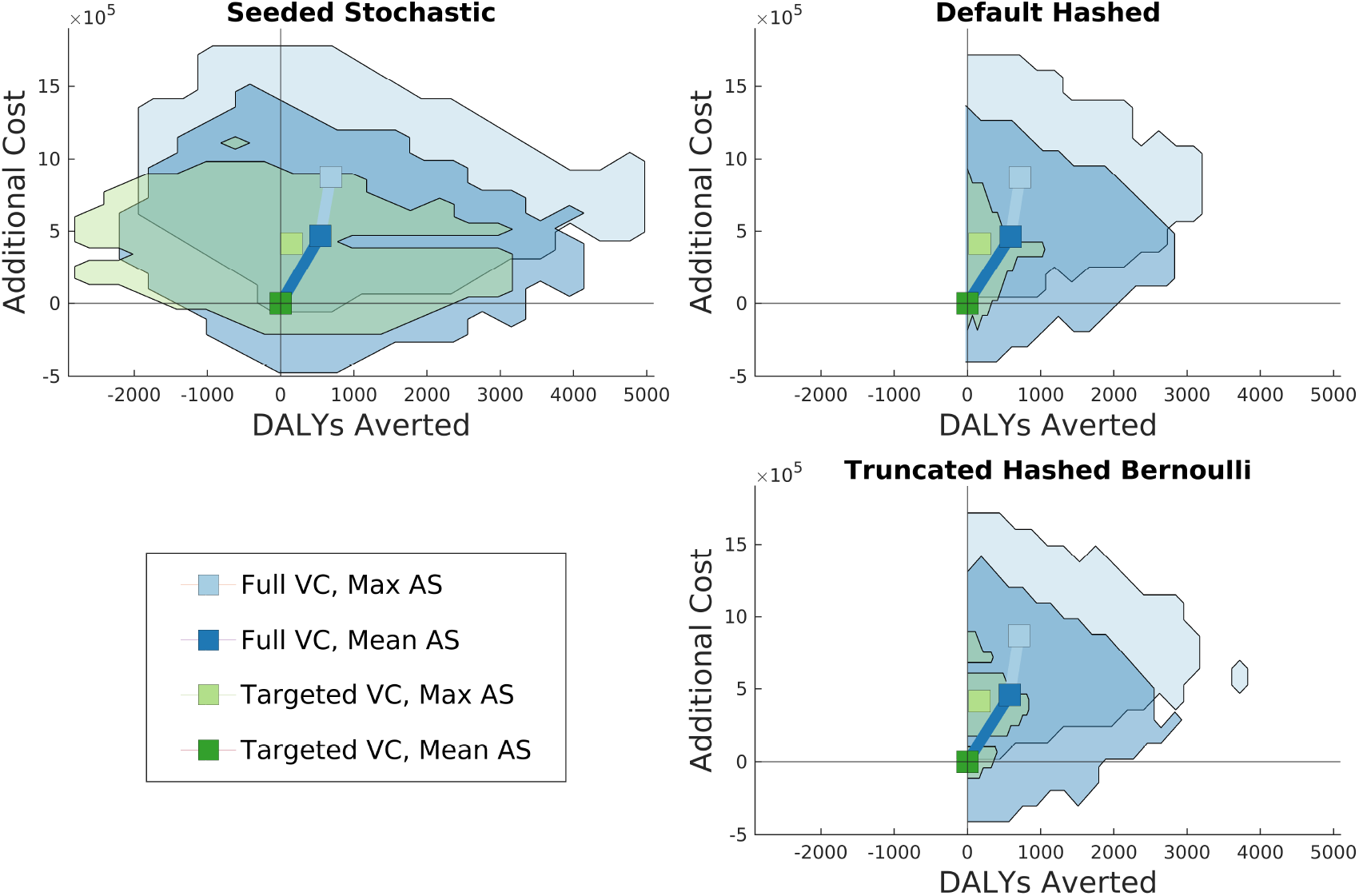
Cost Effectiveness Analysis, showing Costs Averted vs DALYs Averted.

We also note the mean time taken to run a simulation of each of the methods is displayed in Table 3. We note that the new methods do indeed take longer than regular stochastic approaches, however by less than a factor of 2. Additionally, we can see that the truncated Bernoulli takes longer than the default hashing method. When testing alternate maximum numbers of events for the Truncated Bernoulli, we also noted that the larger this maximum event value, the longer each simulation will take—with truncated Bernoulli simulations taking around 2 seconds on average when the maximum number of events was only 20 (as opposed to taking 2.73 seconds when a value of 100 was used).

**Table 3:** Comparison of the computer time taken per simulation of the gHAT model under three different methods. Computer time is in seconds and is given to 3 significant figures.

| Computer time taken per simulation (seconds) |  |  |
| --- | --- | --- |
| Regular Stochastic | Default Hashed | Truncated Bernoulli |
| 1.69 | 2.00 | 2.73 |

Indeed, we would expect the computational time to increase with the maximum number of events, and as such, we opted not to simulate the Bernoulli hashing without truncation.

The change from the seeded simulations to hashing methods is clear in the cost-effectiveness planes (Figure 6). For the default hashing and truncated Bernoulli methods, the bulk of the noise is likely from other sources, such as parameter and cost uncertainty. However, it can be seen when looking at the Targeted VC, Max AS strategy, that we obtain slightly less uncertainty when using the truncated hashed Bernoulli method over the default hashing method. For the hashed-based methods we never find that adding addition interventions (i.e. increasing VC from targeted to full or increasing AS from mean to max) averts fewer DALYs—additional interventions are always beneficial. Using the seeded stochastic method, many of our realisations show worse DALY estimates (negative DALYs averted) for the additional interventions. Finally, looking at the probability of each strategy being optimal (Figure 7), we see that we have more uncertainty under the seeded stochastic method, and so the curves are closer together. The reduction of between-strategy stochastic noise in the hashed-based methods results in much more distinction between the cost-effectiveness acceptability curves and therefore provides a stronger recommendation in which strategy to select at different willingness to pay thresholds.

**Figure 7:**
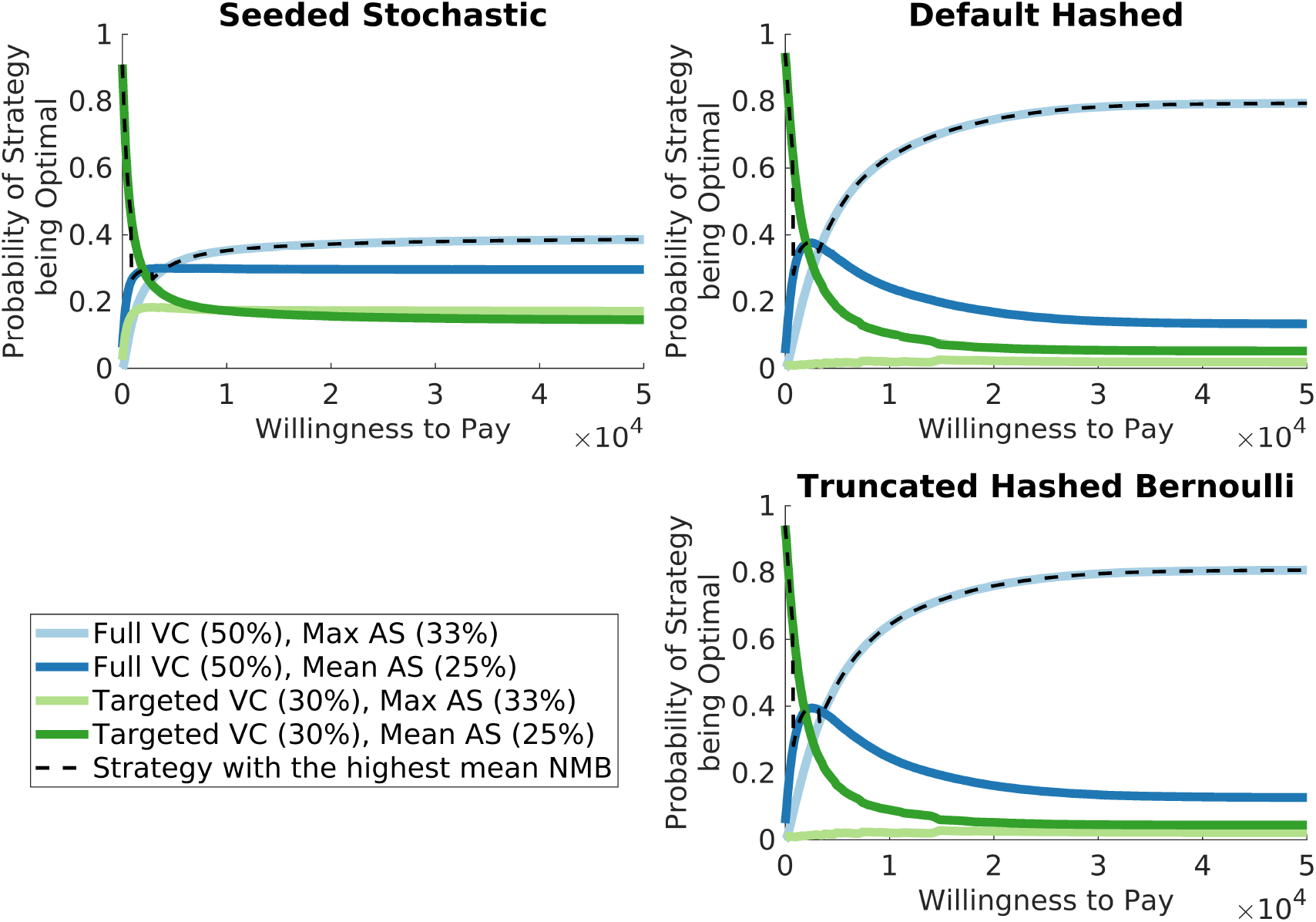
Cost-effectiveness acceptability curves: Probability of each strategy being optimal, versus the willingness to pay to prevent a disability adjusted life year (DALY). VC: vector control; AS: active screening; NMB: net monetary benefit.

## 4 Discussion and Conclusion

### 4.1 Discussion

The SEIRV model was ran using MATLAB [12], and the random number generation was done using the default functions for various distributions. MATLAB is not particularly fast at random number generation and the setting of the random seed when compared to other programming languages. This slow setting of the random seed would explain why it took around four times as long to run the hashed methods compared to the non-hashed ones. Additionally, the slowness of drawing a Poisson random number could explain why the Bernoulli method often appeared to be faster than the default hashing method.

The gHAT model was ran using C++, inside of a MATLAB wrapper, and the random number generation was done using the GSL library, with the taus2 generator. C++ is fast at random number generation, and taus2 is very quick at setting the random seed. This would explain why these methods seemed faster in this model than for the SEIRV model from the previous section, which used MATLAB. Code for all of these models is available at doi.org/10.17605/OSF.IO/U74H3.

The implementation of these methods into both models was not overly complex; however, the gHAT model required more careful consideration than the SEIRV model. The major implementations needed were the creation of the hashing function, the creation of the Bernoulli summation function and the setting of the random seed before event draws. Additionally, for the gHAT model, some GSL [9] functions, such as the binomial and hypergeometric, had to be altered to not use more efficient approaches for some parameter values, as these more efficient approaches often lacked desirable statistical qualities.

Picking a good size of time step for these methods is much the same as picking a good size time step for the regular tau-leap [3], with a notable exception. For the default hashing method, the issue of proportionality causes further issues for larger time steps. This is because longer time steps mean more events are likely to be drawn each step, increasing the impact of this effect. One could also consider implementing an adaptive tau-leaping algorithm, where the size of the time step varies depending on the rates [21].

When originally attempting to implement the Bernoulli method into the gHAT model, it became clear that it would take considerably longer than the other methods (over 100 times longer), and hence the need to implement a truncated version became clear. The truncated version is used when the population of a compartment exceeds 500, and the maximum number of events per time step was set to 100, which given the low rates of the model, was more than sufficient. Originally, a value of 20 was chosen to be the maximum value, but this was too low for the 2000-2025 time period, which had a higher prevalence of gHAT in the population, and so had more events. If prevalence was even higher, or the rates were much larger for some other reason, then the truncated Bernoulli would likely cease to be a reasonable approach, and the default hashing ought to be used instead.

There were also some additional considerations that had to be made regarding the gHAT model. Originally, the number of deaths and the number of passive detections of those in the second infectious stage were drawn as the same event, as they both leave and enter the same compartment (*I*_2_ ↦*R*). For health economic and reporting purposes, the number of deaths and passive detections were needed, so a binomial was then used to determine how many of these events were deaths. Under ordinary simulation methods, this is fine. However, when using reproducible stochastic methods, this could cause issues. In particular, if a counterfactual scenario were considered where the rate of passive screenings was increased. Increasing the level of passive screenings could result in drawing more of the combined event. Depending on the binomial, this could then result in increased deaths, rather than increased passive detections as desired. This was remedied by splitting the passive detecions and the deaths into 2 separate event draws. Those who wish to implement these reproducible methods in their own models should be aware of the possibility of a similar phenomenon occurring.

### 4.2 Conclusion

In this article, we described the default hash-based matching, pseudo-random number generation method for stochastic simulations that was implemented by Pearson and Abbott in the hashprng package [22] to overcome challenges with comparing stochastic model realisations in a fair way. We introduced additional methods building on this hashing idea—namely, the Bernoulli hashing method, and the truncated Bernoulli hashing method which have accuracy and practical advantages. We have discussed the properties of each of these methods for considering counterfactual scenarios and noted how, when compared to other attempts to obtain perfect counterfactuals, they demonstrate advantages in terms of computational complexity and usage on models that are not individual-based.

We note that the default hashing method can be generally applied to any model and will work well, although it has the issue of ‘proportionality’, where the number of events per time step in each compared realisation is going to be from the same percentile of the underlying distribution. The Bernoulli hashing method has the desired statistical properties; however, it should of course be noted that it may be too computationally expensive for larger models. In this case, the truncated Bernoulli hashing method may replace the Bernoulli method for larger models when the rates are low; however, it should be noted that it does lose some of the nicer statistical properties.

We focused on how to apply these approaches to epidemiological models. We first considered a simple example for the implementation of these methods: the SEIRV model (with vaccination). In this, we demonstrated the reduction of noise in between-strategy uncertainty that these various methods result in, which we quantified using the variance in the infections averted by vaccination. We found that in this simple model, whilst both hashing methods provide a large reduction in uncertainty over the regular stochastic method and seeded stochastic method, the Bernoulli hashing method provides a slightly larger reduction without being more computationally expensive.

We then considered its application to the more complicated model of gHAT and its effects on more different metrics. In this analysis, we saw how hashing allowed us to obtain what we believe to be a more accurate demonstration of the probabilities of each strategy being optimal, how it reduced the uncertainty we had on net monetary benefit, and how we had drastically reduced the noise in how much earlier the last transmission event would occur. This exploration allowed us to see how reproducible stochastic modelling would give a fairer comparison of strategies when showing model results to a policy-maker. In doing so, it helps to ensure that additional noise would not negatively impact decisions made by a risk-averse policy-maker. It also demonstrated the usefulness of the truncated Bernoulli hashing method, since the gHAT model is too complex to rapidly simulate with a full Bernoulli hashing method, but has very low rates.

We should note some limitations of these methods. Firstly, they are only applied to tau-leaping simulations, and so may not be ideal for other stochastic modelling approaches. Secondly, applications other than epidemiological models may require other methods or have better model-specific approaches. Additionally, the computational complexity for the Bernoulli methods may be too large to consider using when looking at complex models with high rates. The implementation of this approach can be straightforward for simple models, but more complex models may require deeper thought, as well as greater consideration of how random numbers are drawn. For example, we want to use probability distribution functions that utilise an inverse cumulative distribution function (CDF) from a single uniform random number. Additionally, a random number generator should be used that balances time to set the random seed with desirable statistical properties. Despite these potential complexities, the methods presented in this paper would hopefully be easier to implement than other approaches to simulating counterfactual scenarios.

To summarise, we have shown that hash-based matching, pseudo-random number generation for stochastic simulations can lead to considerable benefit when considering how different strategies compare to one another, and shown when each of the three proposed methods ought to be used.

## Data Availability

The full code used in this paper is available at: http://www.doi.org/10.17605/OSF.IO/U74H3

## Acknowledgements

Thanks to all of the HAT MEPP team and collaborators for the ongoing discussions, which inspired the writing of this paper and for the development of the code that was used to generate the results presented here. The authors would like to thank Dr Erick Mwamba Miaka and the WHO HAT Atlas team for access to historical data from the DRC which allowed for the original gHAT model parameterisation in Antillon et al. [2]. For the purpose of open access, the authors have applied a Creative Commons Attribution (CC-BY) licence to any Author Accepted Manuscript version arising from this submission.

## Ethics approval and consent to participate

This simulation study did not directly use any human case data and used previously published model parameterisation from other publicly available modelling articles. No new data collection took place within the scope of this modelling study.

