## Supplementary Information for "Methods for Reproducible Comparison of Strategies in Stochastic Modelling"

#### 1 Properties of the Truncated Bernoulli

Consider realisation $i$ of strategy A, where we have $N$ (large) susceptibles, and realisation $i$ of strategy B, where we have $N + M$. Under the Bernoulli approach, if we have $k$ infections under strategy A, we must have between $k$ and $k + M$ infections under strategy B. Under the default hashing approach, if we have $k$ infections under strategy A, we must have $\approx k\frac{M+N}{N}$ infections under strategy B. However, in the truncated Bernoulli approach, if we have $k$ infections under strategy A, we must have between $k$ and $m$ infections under strategy B.

This leads to the possible example of $M = 1, k = 0$, that is, for strategy A, we have $N$ susceptibles, and no infection events and for strategy $B$, we have $N + 1$ susceptibles. Under the Bernoulli and default hashing approach, we can only have 0 or 1 infections for strategy B. However, with the truncated hashing approach, we could have between 0 and $m$ infections. Therefore, in incredibly rare circumstances, we can reach our maximum number of infection events by adding an individual to the susceptible population. A more detailed example of this phenomenon can be seen below:

For strategy A, for our population size of $N$, we generate:

$r_{\sigma(m)} \sim Beta(m, N + 1 - m)$ , $\quad R_i \sim U(0, 1)$ , $\quad r_{\sigma(i)} = R_i r_{\sigma(m)}$

For strategy B, for our population size of $N + 1$, we generate:

$\tilde{r}_{\sigma(m)} \sim Beta(m, N + 1 + 1 - m) = r_{\sigma(m)}(1 - \delta)$ , $\quad R_i \sim U(0, 1)$ , $\quad r_{\sigma(i)} = R_i \tilde{r}_{\sigma(m)} = R_i r_{\sigma(m)}(1 - \delta)$,

where $R_i$ and $r_{\sigma(m)}$ are the same as for strategy A, and $\delta$ is some small, positive value.

Let $p$ be the probability of an individual becoming infected. Then, consider the case that for strategy A:

$r_{\sigma(m)} = \frac{p}{1-\delta}$ , $\quad R_i \in (1 - \delta, 1)$

$\implies r_{\sigma(i)} \in (p, \frac{p}{1-\delta}) \quad \implies$ the number of infections under strategy A is 0.

Then, for strategy B, we find that:

$\tilde{r}_{\sigma(m)} = p$ , $\quad R_i \in (1 - \delta, 1)$

$\implies \tilde{r}_{\sigma(i)} \in (p(1 - \delta), p) \quad \implies$ the number of infections under strategy B is $m$.

This means it is possible for a strategy that should always perform worse, to perform better in a given time step — which is not realistic.

### 2 State Based hash

#### 2.1 Theory

The state-based hashing algorithm is an expansion of the default hashing algorithm. Under this approach, the tau-leaping process still draws Poisson random numbers to determine the number of events of each type, and again before each of the random number draws, we shall set the random seed to be the result of a hashing function. However, in this method we vary the hashing function. Notably, the hashing function takes in the state of the system. What this state is can vary per event, but is often going to be the size of the compartment that an individual leaves or joins during the event. For example, infections pass in the exposed class, whilst recoveries pass in the infected class.

#### 2.2 Implementation

The hashing function takes in the time step, system state, salt value, and the event type to produce a hash value. The salt value will be the same between counterfactual realisations if they are in the same state, but unique otherwise. As an example of what this might look like, suppose we have 2 compared strategies — A and B, and the rate of recovery is the same for both. For a realisation of strategy A, we have 10 infected individuals at time $t$, and for B, we have 20. If in A's realisation, we obtain two recoveries, we could obtain any number of recoveries in B's realisation, but the expected number of recoveries in B is still twice that in A. Additionally, if we had the same number of infected individuals under strategies A and B, then we would obtain the same number of recoveries in both of their realisations.

#### 2.3 Properties & Limitations

Under this method, event draws are always independent unless population sizes are the same, which implies the population could behave entirely differently with a change to their size. This additional independence removes the 'proportional' property, but it can cause outcomes to differ more than they would under the default hashing approach.

For our state-based hashing algorithm, we can find it such that the better strategy performs worse in a given timestep. Consider two strategies, A and B, where B results in a lower force of infection. In the first time step, A could have one more infection than B because of this difference. Then, in the next time step, the realisations would have different hash values, as their states differ. Therefore, it is possible for B to have two more infections in this time step than A, out of random chance. This leads to possible future divergence of simulations. In Figure 1 we can see an illustrated example of this phenomenon, which we shall call *stochastic jumping.*

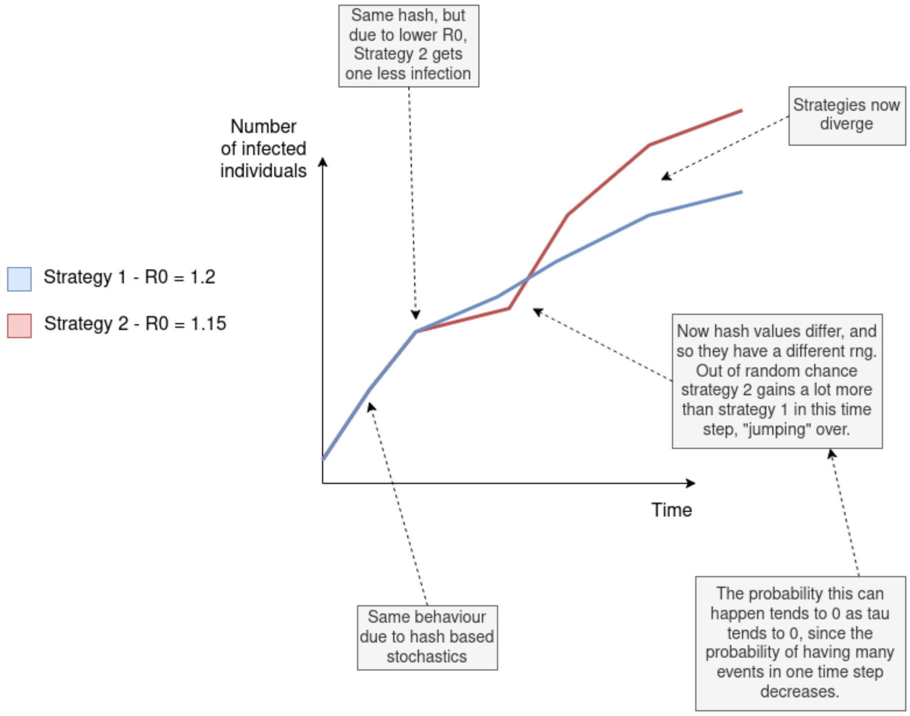


Figure 1: Example of *stochastic jumping* in a generic infection model. Strategy 2 should result in fewer infections.

The lower the value of the time step, the less likely we are to see stochastic jumping as larger draws from the Poisson distributions become less probable, resulting in less frequent diverging of simulations, and so we obtain lower uncertainty.

We can demonstrate this mathematically. Let $X$ be the state of our "worse" strategy, and $Y$ be the state of our "better" strategy. This means:

$$\mathbb{P}(X(t+\tau) = n+k \mid X(t) = k) > \mathbb{P}(Y(t+\tau) = n+k \mid Y(t) = k) \quad \forall n, k > 0. \tag{1}$$

$$\begin{aligned} &\mathbb{P}(\text{Stochastic Jump in a single time step of length } \tau) \\ &= \mathbb{P}(X(t+\tau) < Y(t+\tau) \mid X(t) > Y(t)) \\ &= O(\mathbb{P}(X(t+\tau) = Y(t+\tau) - 1 \mid X(t) = Y(t) + 1)) \\ &= O(\lambda^2 \tau^2 e^{-\lambda\tau}), \\ &\text{for some } \lambda\text{, dependent on the transition rates, where } \exists \lambda_{\min}, \lambda_{\max} \text{ such that} \\ &\qquad \lambda_{\max} > \lambda > \lambda_{\min} \geq 0. \end{aligned} \tag{2}$$

Therefore,

$$\begin{aligned}
&\mathbb{P}(\text{Stochastic jumping in a simulation of length } T \text{ with time step } \tau) \\
&= 1 - \mathbb{P}(\text{No stochastic jumping in a simulation of length } T \text{ with time step } \tau) \\
&= 1 - \prod_{i=1}^{T\tau^{-1}} (1 - \mathbb{P}(\text{Stochastic Jump in time step i, of length } \tau)) \\
&= O\left(1 - \prod_{i=1}^{T\tau^{-1}} (1 - \tau^2 e^{-\lambda_i \tau})\right) \\
&= O\left(1 - (1 - \lambda_{\max}\tau^2 e^{-\lambda_{\min}\tau})^{T\tau^{-1}}\right) \\
&= O\left(T\tau^{-1}\lambda_{\max}\tau^2 e^{-\lambda_{\min}\tau}\right) \\
&= O(\tau e^{-\lambda_{\min}\tau})
\end{aligned} \tag{3}$$

Finally, we can see that:

$$\lim_{\tau \to 0} \ \mathbb{P}(\text{Stochastic jumping in an entire simulation with time step } \tau) \ = \ 0 \tag{4}$$

Note: the big $O$ notation used above is in the limit as $\tau \to 0$.

#### 2.4 Comparison to other methods

| | **Default Hashing** | **State-based Hashing** | **Hashed Bernoulli** | **Truncated Bernoulli** |
|---|---|---|---|---|
| Does the hash function depend on the state? | No | Yes | No | No |
| Distribution of number of events | Poisson | Poisson | $\sum$ Bernouuli | $\sum$ Bernoulli |
| Dependence on the number of events drawn between realisations | Always dependent and 'proportional' | Only when at the same state and time | Dependent between realisations but not 'proportional' | Dependent between realisations, but underlying beta and uniform distributions are 'proportional'. |
| Can worse strategies perform better in a given time step | No | Yes, but the probability of this tends to 0 as the time step tends to 0 | No | Yes, with very low probability, which tends to 0 as the time step tends to 0. |
| Computational speed | Quick | Quick | Slow (scales with population size) | Quick (scales with truncation point) |

Table 1: Comparison of the various methods

### 3 Hashing Algorithm and Random Number Generation

#### 3.1 Random number generation

When drawing random numbers, we want a proportionality between 2 compared realisations at the same random state. For example, let's consider we have 2 compared realisations, both with 999 susceptible individuals at time 0. Assume strategy 1 results in an infective rate approximately twice that of strategy 2. ($R_0 = 4$ vs $R_0 = 2$) Then, if a realisation of strategy 1 has an infection in the first time step, strategy 2 should have either 0 or 1.

Additionally, if there are any other random events occurring, these should likely also have a similar random effect. (e.g. if sampling the population for screening, should not find more infections for a strategy with fewer infected individuals). The setting of a random seed alone might not be sufficient to do this, depending on the method of random number generation. This is due to functions for drawing from distributions often using approaches to increase computational speed.

An inverse cumulative density function method will always give the desired random properties, but users should be careful about the possibility of other methods resulting in unwanted outcomes. It should also be noted that an inverse cumulative density function may be computationally expensive compared to other approaches that still give the desired behaviour.

#### 3.2 Using the random hash

```
calculate_hash(int const &t, int const &Salt, int const &event_num)
{
    // Hashing function
    // Three large prime numbers for hashing
    unsigned long prime1 = 98610401689;
    unsigned long prime2 = 3086358019;
    unsigned long prime3 = 2147483647;

    unsigned long myhash = (Salt * prime1) ^ ((t+999) * prime3);
    myhash = (myhash + (event_num * prime2)) ^ ((myhash >> 6));
    myhash = myhash + 1;
    // ensure we return a non-zero value
    return myhash;
}
}
```

This is our hashing algorithm.

Before each random number draw, the random seed must be set to the result of the hashing algorithm as follows:

```
rng(calculate_hash(t,Salt, event_num))
```

$t$ is the time value. It can be any measurement of time (Year, Day, timestep number, etc.), so long as it changes every time step.

Salt is a sort of base random seed that is the same for each compared realisation. The realisation number could be used here.

event num is a number that uniquely describes each event. e.g. 1 for infection, 2 for recovery, etc. This ensures different events don't have the same random seed.

##### 3.2.1 Hashing function under the state-based hash

Under the state-based hashing method, we must alter the hashing function to include the state of the system. As such, our function now looks like:

```
uint64_t calculate_hash(double const &t, int const &Salt, double const &system_state_hashed,
double const &event_num)
{
    uint64_t prime1 = 98610401689; //Five large prime numbers for hashing
    uint64_t prime2 = 16860199001;
    uint64_t prime3 = 2147483647;
    uint64_t prime4 = 54771844201;
    uint64_t prime5 = 3086358019;

    uint64_t myhash = uint64_t(Salt * prime1 + system_state_hashed * prime2) ^
                    uint64_t((t+999) * prime3);
    myhash = uint64_t(myhash + (system_state_hashed * prime4) + (event_num * prime5)) ^
                    uint64_t((myhash >> 6));
    return myhash;
}
```

Again, before each random number draw, the random seed must be set to the result of the hashing algorithm as follows:

```
rng(calculate_hash(t,Salt, system_state_hashed, event_num))
```

$t$, Salt and event num are the same as before.

system state hashed is the value of the state being used to set the randomness. This should always be the state that is being left, with the exception of infectious events, where this should be the class that newly infected individuals end up in.

### 4 SEIRV model

#### 4.1 Model explanation - further details

Our SEIRV model is a simple compartmental model based on the ubiquitous SEIR model presented in the literature (e.g. see Keeling and Rohani, [?]).

- The susceptible class (S) are those who do not have the disease but could get it in the future.
- The exposed class (E) are those who have gotten the disease but are not currently infectious.
- The infected class (I) are those who have gotten the disease and are currently infectious.
- The recovered class (R) are those who had the disease and are no longer infectious and cannot be reinfected.
- The vaccinated class (V) are those who got vaccinated before getting infection, and as such cannot become infected. Vaccination occurs at a constant rate from the susceptible class S.

The following events can happen in the model:

- An exposure event. A susceptible becomes exposed to the disease. This happens at rate $\frac{\beta I}{N}$.
- An infectious event. An exposed individual becomes infected. This happens at rate $\sigma$.
- A recovery event. An infected individual recovers from the disease. This happens at rate $\gamma$.
- A vaccination event. A susceptible individual is vaccinated from the disease. This happens at rate $\alpha$.

This can also be represented as a system of ODEs, which is what we use for the deterministic model:

$$\begin{aligned}\frac{\mathrm{d}S}{\mathrm{d}t} &= -\beta SI - \alpha S \\ \frac{\mathrm{d}E}{\mathrm{d}t} &= \beta SI - \sigma E \\ \frac{\mathrm{d}I}{\mathrm{d}t} &= \sigma E - \gamma I \\ \frac{\mathrm{d}R}{\mathrm{d}t} &= \gamma I \\ \frac{\mathrm{d}V}{\mathrm{d}t} &= \alpha S\end{aligned} \tag{5}$$

We will give the infection parameters the following distributions:

- $\beta \sim \Gamma(4, 0.25)$
- $\sigma \sim \Gamma(10, 0.1)$
- $\gamma \sim \Gamma(3, 0.15)$

Meanwhile, the vaccination parameter $\alpha$ will be something we will vary to simulate different levels of intervention.

##### 4.1.1 State-based set up

The following is the list of states used in the hashing function for each event.

- For the exposure event, we use the exposed class.
- For the infectious event, we use the exposed class.
- For the recovery event, we use the infected class.
- For the vaccination event, we use the susceptible class.

#### 4.2 Additional Results

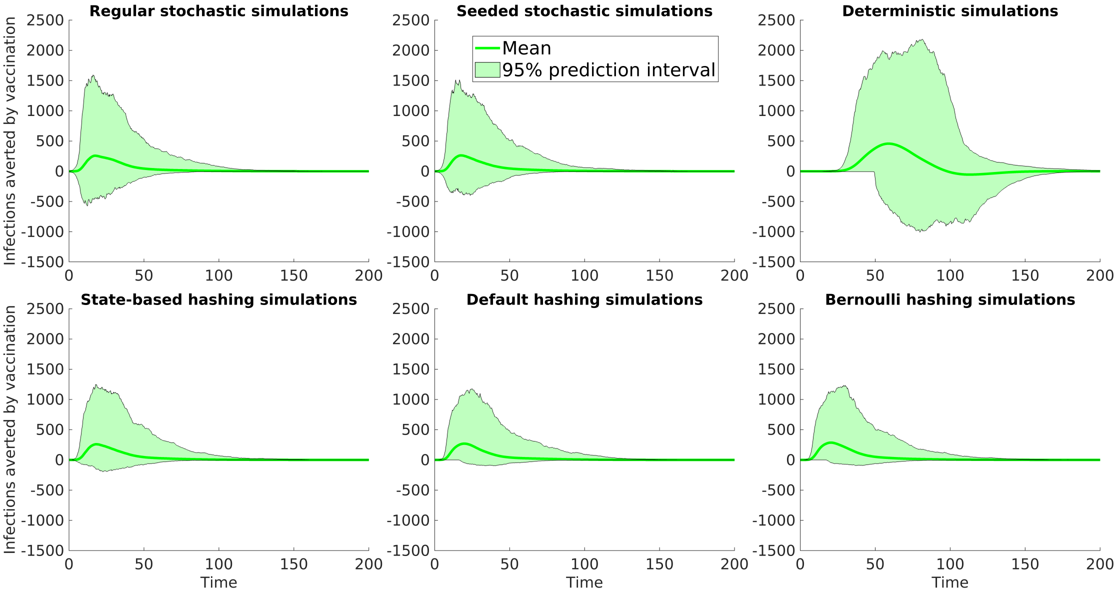


Figure 2: Plots of the infections averted by undertaking vaccination, where vaccination rate is 0.008 day$^{-1}$, for our 6 different simulation methods. Stochastic simulations had a tau leap of 0.25 for these results.

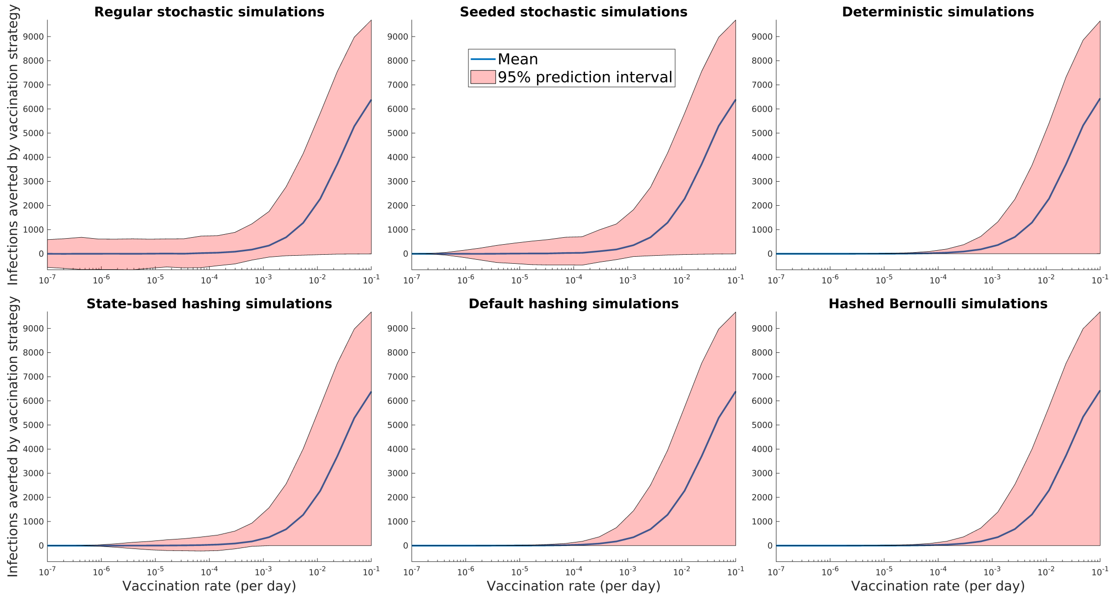


Figure 3: Plots of the infections averted by undertaking vaccination, where vaccination rate is varied, for our 6 different simulation methods. Stochastic simulations had a tau leap of 0.04 for these results.

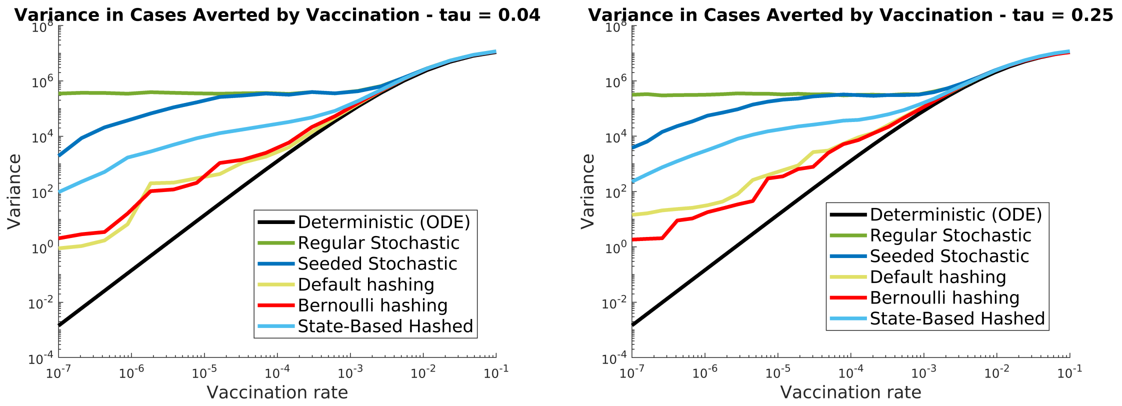


(a) Time step $\tau = 0.04$

(b) Time step $\tau = 0.25$

Figure 4: Variance in the number of infections averted for different simulation methods, against the vaccination rate. A smaller and a larger time step are used.

### 5 gHAT model

#### 5.1 Model explanation

The gHAT model presented here has been extensively used by the Warwick "HAT MEPP" team for modelling transmission of the parasite *Trypanosoma brucei gambiense* between humans and tsetse in different settings. There have been numerous improvements to the model since its original publication [**?**]; the version used here mimics that used by Antillon et al. [2] for modelling of gHAT in the Democratic Republic of Congo. Although all information is provided in this SI and the code to make this analysis reproducible, for justification of modelling and intervention assumptions, the reader should refer to the SI of Antillon et al. [2].

The following is a list of all the compartments in the model. This list is split into tsetse and human components. It should also be noted that each human component has a low- and high-risk group.

- $P_V$, the tsetse pupal class
- $S_V$, the tsetse susceptible, teneral class. Tsetse are teneral before their first blood meal, where they have a higher susceptibility to becoming infected.
- $E_{1V}$, the tsetse exposed stage 1 class
- $E_{2V}$, the tsetse exposed stage 2 class
- $E_{3V}$, the tsetse exposed stage 3 class. Note that we use the linear chain trick: having 3 exposed classes to create an Erlang distributed extrinsic incubation period.
- $I_V$, the tsetse infectious class
- $G_V$, the tsetse susceptible, non-teneral class
- $S_H$, the human susceptible class

- $E_H$, the human exposed class
- $I_{1H}$, the human infectious stage 1 class
- $I_{2H}$, the human infectious stage 2 class
- $R_H$, the human recovered class

We note that the code contains options for skin-only parasite infection [3], but we omit this from our model.

The following is a list of each event that can occur within the stochastic tau-leaping gHAT model. For each of these we will state the transition that occurs, the rate at which it occurs, and the compartment to be included in the hashing along with the Salt value and the time, if the state-based hash is used.

- New Infection Event
  - Transition: $S_H \to E_H$
  - Rate: $I_V \alpha m_{\text{eff}} f \frac{1}{N_H}$
  - Compartment for hashing: $S_H$, the human susceptible class
- End Latency Period Event
  - Transition: $E_H \to I_{1H}$
  - Rate: $\sigma p_{BS}$
  - Compartment for hashing: $E_H$, the human exposed class
- Die naturally in exposed state
  - Transition: $E_H \to S_H$
  - Rate: $\mu$
  - Compartment for hashing: $E_H$, the human exposed class
- Progress to stage 2 of infection
  - Transition: $I_{1H} \to I_{2H}$
  - Rate: $\phi$
  - Compartment for hashing: $I_{1H}$, the human infectious stage 1 class
- Die naturally in stage 1
  - Transition: $I_{1H} \to S_H$
  - Rate: $\mu + \omega_{IB}$
  - Compartment for hashing: $I_{1H}$, the human infectious stage 1 class
- Passive detection from stage 1
  - Transition: $I_{1H} \to R_H$

- Rate: $\eta$
- Compartment for hashing: $I_{1H}$, the human infectious stage 1 class

- Die naturally in stage 2
  - Transition: $I_{2H} \to S_H$
  - Rate: $\mu$
  - Compartment for hashing: $I_{2H}$, the human infectious stage 2 class

- Die naturally or lose immunity in recovered
  - Transition: $R_H \to S_H$
  - Rate: $\omega + \mu$
  - Compartment for hashing: $R_H$, the human recovered class

- Passive detection or death from stage 2
  - Transition: $I_{2H} \to R_H$
  - Rate: $\gamma$
  - Compartment for hashing: $I_{2H}$, the human infectious stage 2 class

It should be noted that the tsetse dynamics are simulated with an ODE model, as there are large numbers of events in a short time period. This is done with a Runge-Kutta approach to approximate the daily ODE tsetse dynamics[5]. The tsetse model is as follows:

$$
\begin{aligned}
\frac{\mathrm{d}P_V}{\mathrm{d}t} &= B_V N_H - (\xi_V + \tfrac{P_V}{K})P_V \\
\frac{\mathrm{d}S_V}{\mathrm{d}t} &= \xi_V \mathbb{P}(\text{pupating})P_V - \alpha S_V - \mu_V S_V \\
\frac{\mathrm{d}E_{1V}}{\mathrm{d}t} &= \alpha\big(1 - f_T(t)\big)p_V\big(\textstyle\sum_i f_i \dfrac{(I_{1Hi} + I_{2Hi})}{N_{Hi}}\big)(S_V + \varepsilon G_V) \\
&\quad -(3\sigma_V + \mu_V + \alpha f_T(t))E_{1V} \\
\frac{\mathrm{d}E_{2V}}{\mathrm{d}t} &= 3\sigma_V E_{1V} - (3\sigma_V + \mu_V + \alpha f_T(t))E_{2V} \\
\frac{\mathrm{d}E_{3V}}{\mathrm{d}t} &= 3\sigma_V E_{2V} - (3\sigma_V + \mu_V + \alpha f_T(t))E_{3V} \\
\frac{\mathrm{d}I_V}{\mathrm{d}t} &= 3\sigma_V E_{3V} - (\mu_V + \alpha f_T(t))I_V \\
\frac{\mathrm{d}G_V}{\mathrm{d}t} &= \alpha(1 - f_T(t))\Big(1 - p_V\big(\textstyle\sum_i f_i \dfrac{(I_{1Hi} + I_{2Hi})}{N_{Hi}}\big)\Big)S_V \\
&\quad -\alpha\Big(f_T(t) + \big(1 - f_T(t)\big)p_V\varepsilon\big(\textstyle\sum_i f_i \dfrac{(I_{1Hi} + I_{2Hi})}{N_{Hi}}\big)\Big)G_V \\
&\quad -\mu_V G_V
\end{aligned}
\tag{6}
$$

$$
f_T(t) = p_{\text{target die}}\left(1 - \frac{1}{1 + \exp(-0.068(\text{mod}(t, 182.5) - 127.75))}\right)
$$

The human compartments also have the option to be simulated using an ODE model, but we do not use this in our results. The equations for the human compartments is:

$$\begin{aligned}
\frac{\mathrm{d}S_{Hi}}{\mathrm{d}t} &= \mu_H N_{Hi} + \omega_H R_{Hi} - \alpha m_{\text{eff}} f_i \frac{S_{Hi}}{N_{Hi}} I_V - \mu_H S_{Hi} \\
\frac{\mathrm{d}E_{Hi}}{\mathrm{d}t} &= \alpha m_{\text{eff}} f_i \frac{S_{Hi}}{N_{Hi}} I_V - (\sigma_H + \mu_H) E_{Hi} \\
\frac{\mathrm{d}I_{1Hi}}{\mathrm{d}t} &= \sigma_H E_{Hi} - (\varphi_H + \eta_H(Y) + \mu_H) I_{1Hi} \\
\frac{\mathrm{d}I_{2Hi}}{\mathrm{d}t} &= \varphi_H I_{1Hi} - (\gamma_H(Y) + \mu_H) I_{2Hi} \\
\frac{\mathrm{d}R_{Hi}}{\mathrm{d}t} &= \eta_H(Y) I_{1Hi} + \gamma_H(Y) I_{2Hi} - (\omega_H + \mu_H) R_{Hi}
\end{aligned} \tag{7}$$

##### 5.1.1 Parameterisation

The fitted parameters [1] seen in Table 2 generally correspond to assumed biological values that are not expected to vary in different locations. The human population is taken to be 180,000 for our example region. The fitted parameters in Table 3 come from model fitting work [1] done with MCMC to find posteriors; however, we will be using these as illustrative parameterisations and our simulations do not use any real data pertaining to the past or planned activities of any particular health zone. The tsetse to human relative density is calculated from $R_0$[4]. The base probability of vector control killing a tsetse when it takes its bloodmeal, $p_{\text{target die}}$, is chosen to get the desired level of reduction in the tsetse population (which depends on the strategy).

| Notation | Description | Value |
|---|---|---|
| $N_H$ | Total human population size | 180,000 |
| $\mu_H$ | Natural human mortality rate | $5.4795 \times 10^{-5}\text{days}^{-1}$ |
| $B_H$ | Total human birth rate | $= \mu_H N_H$ |
| $\sigma_H$ | Human incubation rate | $0.0833\text{days}^{-1}$ |
| $\varphi_H$ | Stage 1 to 2 progression rate | $0.0019\text{days}^{-1}$ |
| $\omega_H$ | Recovery rate | $0.006\text{days}^{-1}$ |
| $\xi_V$ | Pupal death rate | $0.037\text{days}^{-1}$ |
| $K$ | Pupal carrying capacity | $= 111.09 N_H$ |
| $\mathbf{P}$(pupating) | Probability of pupating | 0.75 |
| $\mu_V$ | Tsetse mortality rate | $0.03\text{days}^{-1}$ |
| $\sigma_V$ | Tsetse incubation rate | $0.034\text{days}^{-1}$ |
| $\alpha$ | Tsetse bite rate | $0.333\text{days}^{-1}$ |
| $B_V$ | Tsetse birth rate | $0.0505\text{days}^{-1}$ |
| $p_V$ | Probability tsetse infected per infectious bite | 0.065 |
| $\epsilon$ | Reduced non-teneral susceptibility factor | 0.05 |
| $f_H$ | Proportion of blood-meals on humans | 0.09 |
| $m_{\text{eff}}$ | Probability human infected per infectious bite | Fitted(?) |

Table 2: Fixed parameters of the gHAT model. Adapted from Antillon et al[1]

| Notation | Description | Percentiles of Prior Distribution [2.5, 50 & 97.5%] |
|---|---|---|
| $m_{\text{eff}}$ | Tsetse to human relative density | Calculated from $R_0$ |
| $R_0$ | Basic reproduction number (NGM approach) | [1.003, 1.069, 1.369] |
| $\eta_H(Y)$ | Stage 1 treatment rate | [4.59, 17.1, 42.9] $\times 10^{-5}$ |
| $\gamma_H(Y)$ | Stage 2 treatment rate | [7.59, 40.7, 121] $\times 10^{-4}$ |

Table 3: Fitted parameters of the gHAT model. Adapted from Antillon et al [1] [4]

| 2000 | 2001 | 2002 | 2003 | 2004 | 2005 | 2006 | 2007 | 2008 | 2009 | 2010 | 2011 |
|---|---|---|---|---|---|---|---|---|---|---|---|
| 21622 | 27943 | 23113 | 25286 | 21657 | 26020 | 22630 | 26541 | 26893 | 27482 | 24506 | 20839 |

| 2012 | 2013 | 2014 | 2015 | 2016 | 2017 | 2018 | 2019 | 2020 | 2021 | 2022 |
|---|---|---|---|---|---|---|---|---|---|---|
| 22290 | 29134 | 21524 | 56517 | 50767 | 59923 | 41564 | 48854 | 42134 | 59238 | 40093 |

Table 4: Historical Active Screening Levels

#### 5.2 Modelling interventions (past and future)

We assume that passive screening interventions are always ongoing, at rates outlined in the parameter table. For active screening, we assume that the number of people screened historically from the years 2000-2022 is as seen in Table 4. In 2023 and 2024 the level of active screening is mean, and then onwards, the level of AS is either done at the mean level or the max level. At the mean level, the mean of the years 2018-2022 is used for future screening, whilst at the max level, the maximum value of the years 2000-2022 is used.

For vector control, this does not begin until 2025. From 2025 onwards, vector control is deployed twice a year. The % reduction in tsetse after the first year is 30% under a targeted approach, and is 50% under a full approach. We consider a 30 year time horizon, and active screening and vector control interventions are stopped after 3 consecutive years of no case reporting.

#### 5.3 Evaluating disease burden

We can calculate the disability-adjusted life years (DALYs) incurred from gHAT infection under a strategy. Using the model, the weighted person-years spent in each stage of infection can be calculated and the total number of deaths can be multiplied by the average years of life lost to gHAT to obtain the total person-years lost to death. The disability weightings for gHAT infection are assumed to be 0.14 during the first stage of infection and 0.54 during the second stage [2]. The years lost to death is the life expectancy, 60.02, minus the average age at death of 26.63 [1], which, with the yearly discounting of 3%, becomes 21.54.

$\text{DALYS} = 0.14 \times \text{PersonYears}_1 + 0.54 \times \text{PersonYears}_2 + 21.54 \times \text{Deaths}$

#### 5.4 The cost function

We can also determine the costs of performing an intervention. We define a cost function similar to that used in Antillon et al. [2], with some small changes, simplifying the screening costs, and removing the cost of monitoring tsetse from the vector control costs. For a year, $t$, the cost for a strategy $S$ in 2022 USD is:

$$
\begin{aligned}
\text{Cost}_t = &\text{Fixed AS Costs} * \mathbb{1}\{\text{Is screening in year t when doing strategy S}\}\\
&+\text{Testing Costs} * \text{People Screened in year t when doing strategy S}\\
&+\text{Confirmation Cost} * \text{Infected People Screened in year t when doing strategy S}\\
&+\text{Fixed VC Cost} * \mathbb{1}\{\text{Is doing VC in year t when doing strategy S}\}\\
&+\text{VC cost per linear km} * \text{linear km of river under strategy S}
\end{aligned} \tag{8}
$$

The linear km of rivers is specific to each health region, and for our example region, we have used a value of 200km when doing targeted VC, and 600km when doing full VC.

The rest of these are costs, each of which has a distribution taken from the literature. These are:

- Fixed AS Costs $\sim \Gamma(56.18, 1412.88) + \Gamma(25.31, 747.18)$

These are the management costs of active screening, plus the cost of an AS team, both taken from [2].

- Testing Costs $\sim \Gamma(12.11, 0.1319)$

This is the cost of a CATT test, taken from [2].

- Confirmation Cost $\sim \Gamma(8.47, 1.36)$

This is the cost of confirming a case of gHAT through microscopy, taken from [2].

- Fixed VC Cost $\sim \Gamma(8.47, \frac{8.47}{\mu})$
where $\mu = 9616.5 * 1.2194$

For this, we assume the fixed VC cost is the cost of travel. We also scale by 1.2194 to account for inflation.

These values are taken from the Yasa Bonga VC costs [6] and scaled accordingly.

As we only have 1 set of data to determine these costs (from Yasa Bonga), we take a gamma distributed cost, with the mean taken from these data and the 95% CI set to be 0.5 and 2 times the mean.

- VC cost per linear km $\sim \Gamma(8.47, \frac{8.47}{\mu})$
where $\mu = \frac{43340.5*1.2194}{417}$

For this, we assumed that all other non-monitoring VC costs scale linearly with the extent (in linear km) of the VC. We also scale by 1.2194 to account for inflation.

These values are taken from the Yasa Bonga VC costs [6] and scaled accordingly.

As we only have 1 set of data to determine these costs (from Yasa Bonga), we take a gamma distributed cost, with the mean taken from these data and the 95% CI set to be 0.5 and 2 times the mean.

#### 5.5 Cost effectiveness analysis

It should be noted that both DALYs and Costs are given a 3% yearly discounting. With these DALYs and Costs, we can then perform a cost-effectiveness analysis (CEA). One way we can do this is by calculating the net monetary benefit (NMB). We require a willingness to pay (WTP) value of the policymaker, which is how much they are willing to pay to prevent a DALY. Then, for each strategy, when compared to a comparator strategy:

$$\text{NMB} = \text{Costs Averted} + \text{DALYs Averted} \times \text{WTP} \tag{9}$$

Another approach we can use is to plot a cost-effectiveness plane of DALYs averted and Costs averted when compared to the comparator strategy (the one with the lowest cost). We can then calculate the incremental cost-effectiveness ratio values for each strategy. The following is an algorithm for determining these:

1. Order all considered strategies by their cost: Strategy 1, Strategy 2, ...
2. Remove any strategy that results in more (or equal) DALYs than the strategy before it. These are dominated.
3. Calculate the ICER for each strategy (except Strategy 1, which has a value of 0).
   (a) $\text{ICER} = \frac{C_i - C_{i-1}}{D_{i-1} - D_i}$
   (b) $C_i$ is the cost of strategy $i$, and $D_i$ is the DALYs under strategy $i$.
4. Remove any strategy that has a higher ICER than the strategy after it. These are weakly dominated.
5. Repeat back to step 1 until all strategies remaining (non-dominated) are ordered such that their ICER values are ascending.

Dominated strategies are not economically optimal. Non-dominated strategies are economically optimal when their ICER value is the largest of all strategies with ICER values less than the willingness to pay threshold.

#### 5.6 Additional Results

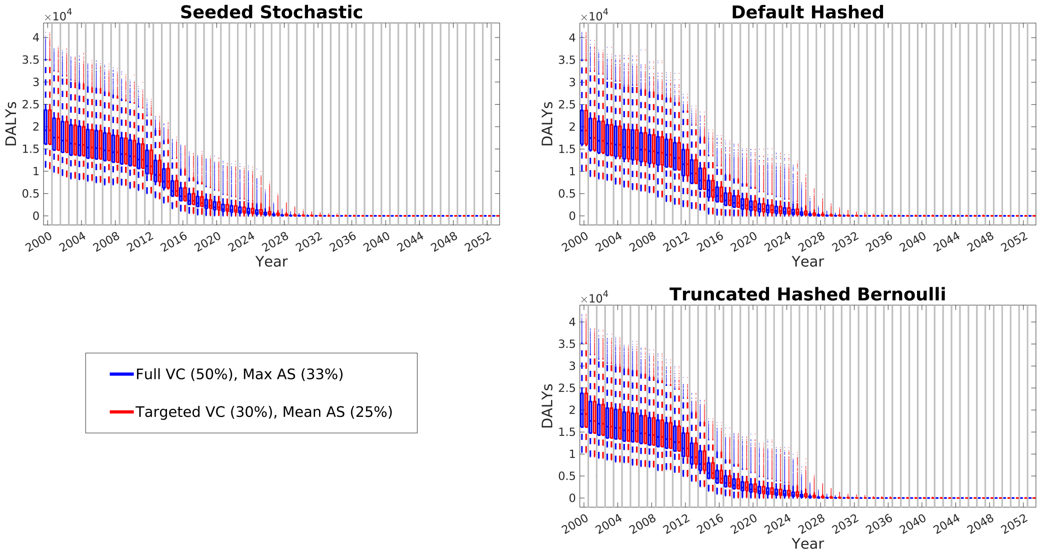


Figure 5: DALYs each year, for each simulation method. We note that these are expected to be similar (and are) across the different realisation methods. We also note how the impact of the differing strategies can be seen from 2025 onwards.

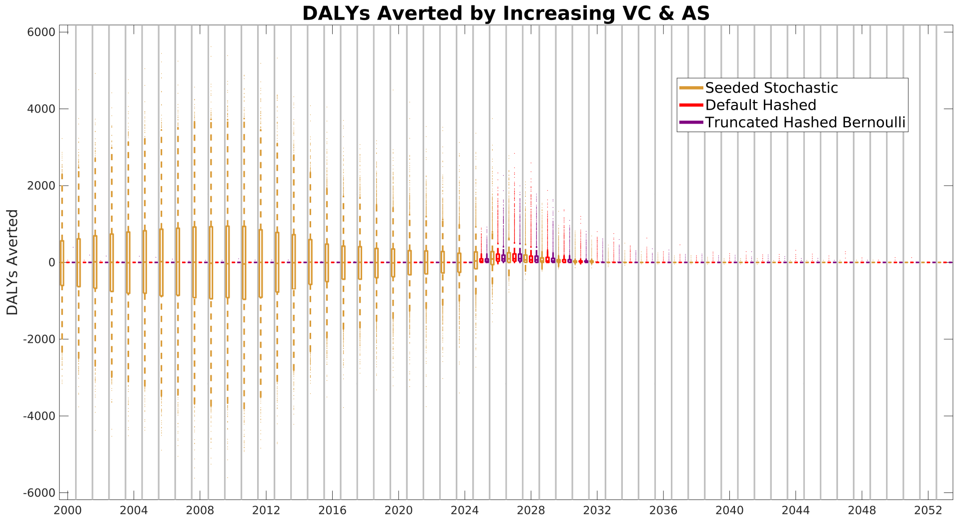


Figure 6: DALYs Averted each year by going from Mean AS and Targeted VC to Max AS and Full VC, for different simulation methods. We note before 2025, the hashing methods ensure DALYs match. We also note that after 2025, hashing methods keep the DALYs averted positive.

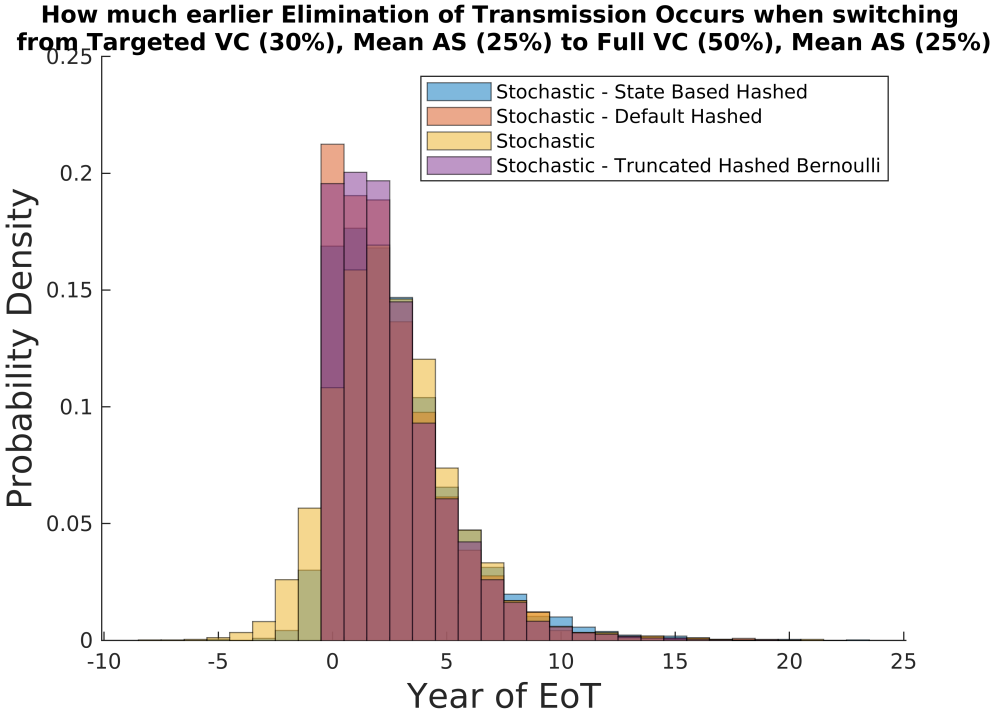


Figure 7: How much earlier Elimination of Transmission occurs for the different methods

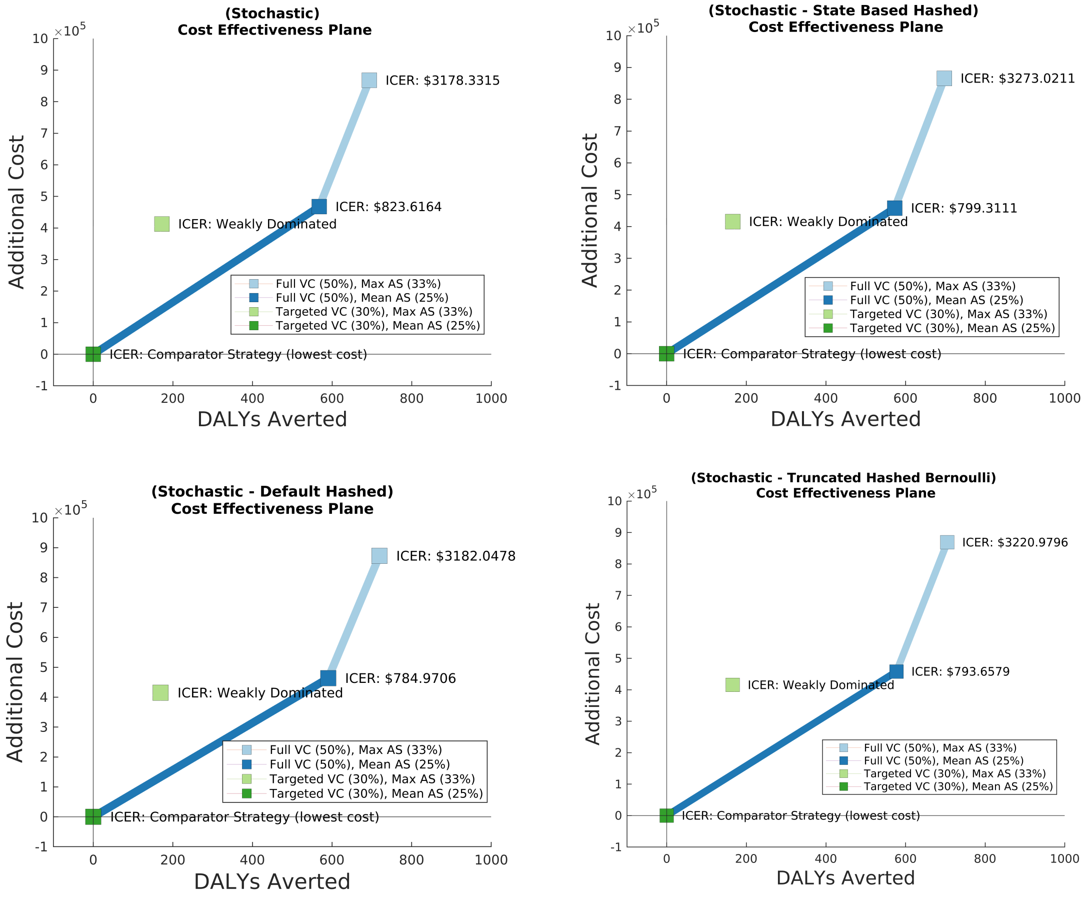


Figure 8: Zoomed in Cost Effectiveness Analysis, showing only the mean values.

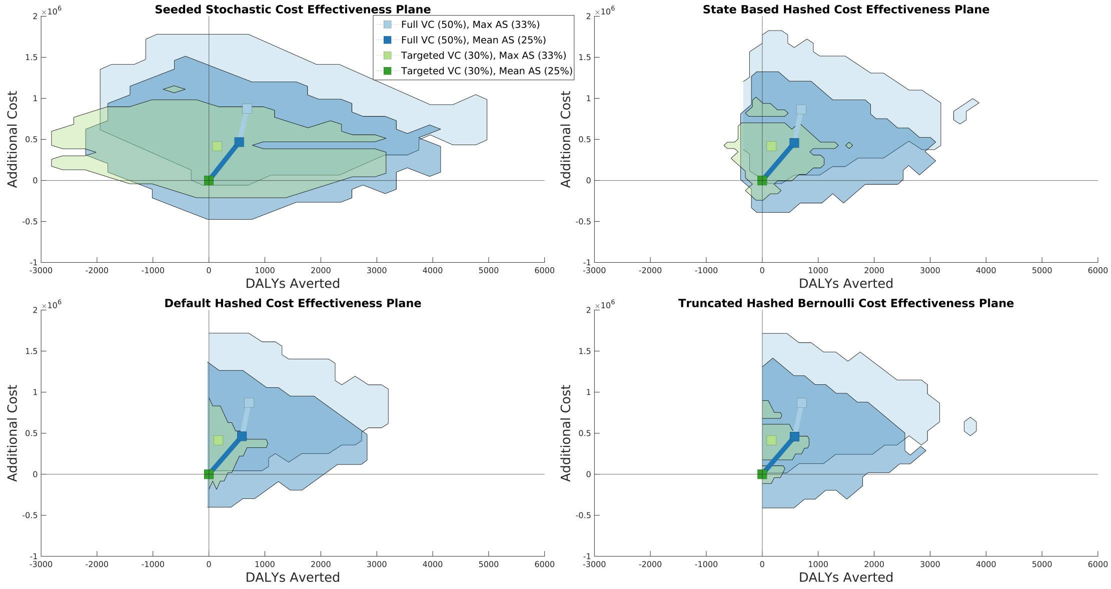


Figure 9: Cost Effectiveness Analysis, showing Costs Averted vs DALYs Averted.

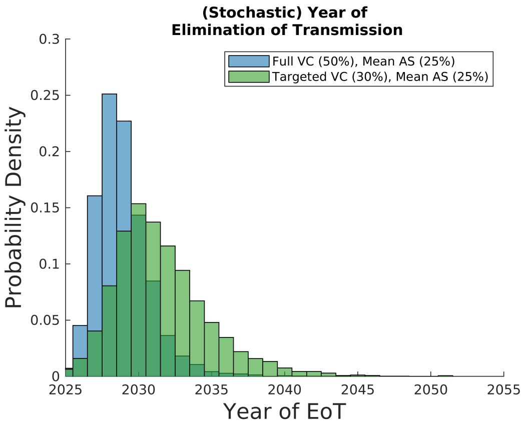


(a) Stochastic Simulations

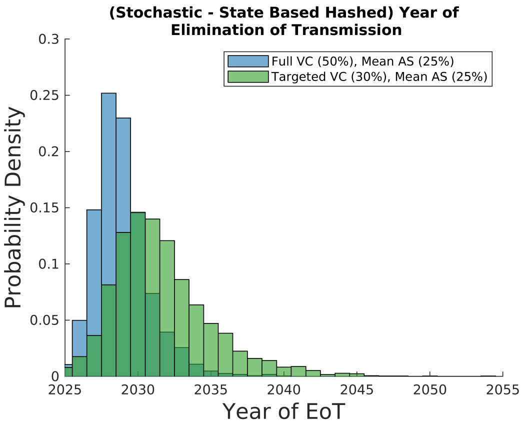


(b) State-Based Hashed Stochastic Simulations

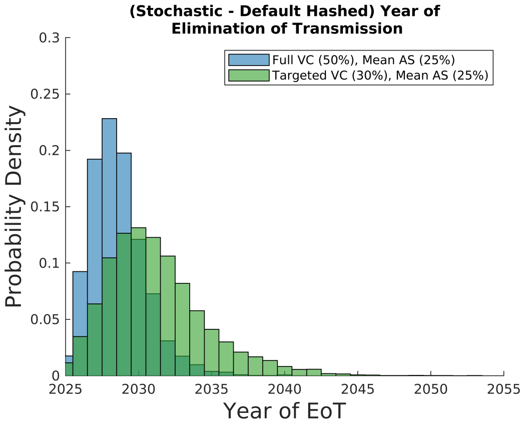


(c) Default Hashed Stochastic Simulations

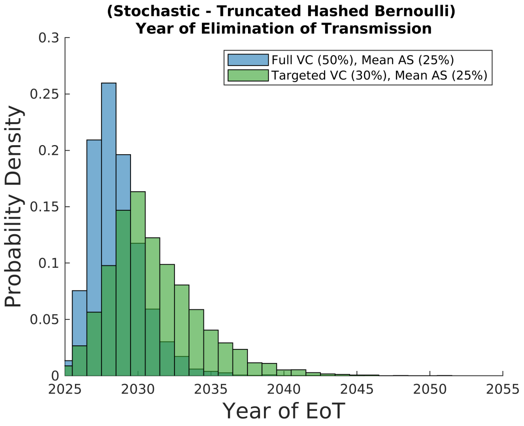


(d) Truncated Hashed Bernoulli Stochastic Simulations

Figure 10: Elimination of Transmission for 2 different strategies.

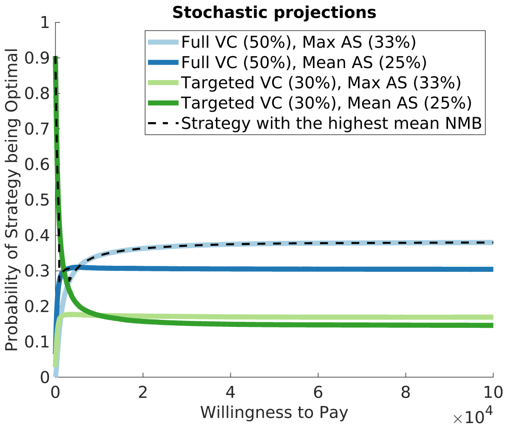


(a) Stochastic Simulations

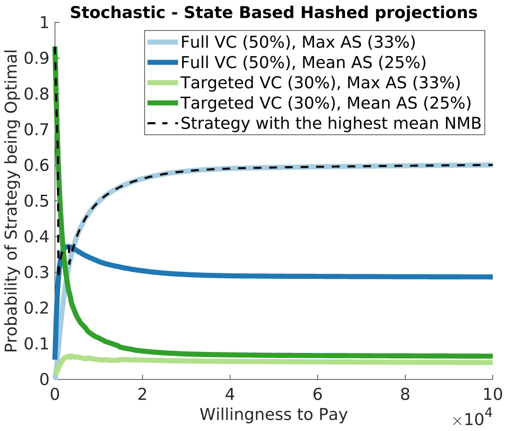


(b) State-Based Hashed Stochastic Simulations

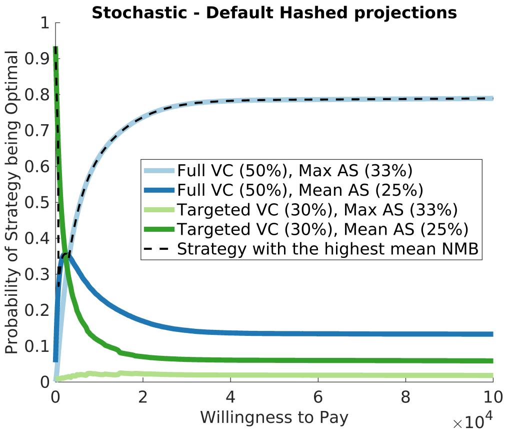


(c) Default Hashed Stochastic Simulations

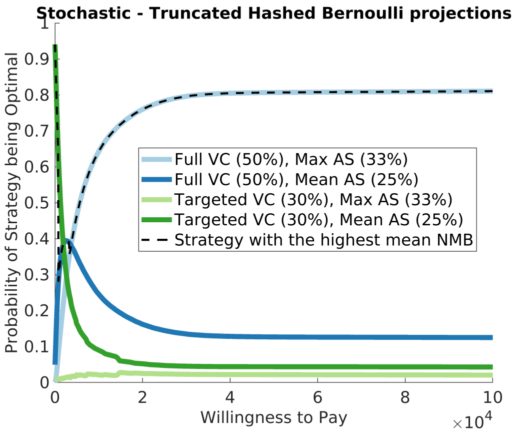


(d) Truncated Hashed Bernoulli Stochastic Simulations

Figure 11: Probability of each strategy being optimal, versus the Willingness to Pay to prevent a DALY.

#### References

[1] Marina Antillon, Ching-I Huang, Ronald E. Crump, Paul E Brown, Rian Snijders, Erick Mwamba Miaka, Matt J. Keeling, Kat S. Rock, and Fabrizio Tediosi. Cost-effectiveness of sleeping sickness elimination campaigns in five settings of the Democratic Republic of Congo. *Nat Commun*, 2022.

[2] Marina Antillon, Ching-I Huang, Samuel A. Sutherland, Ronald E. Crump, Paul E. Brown, Paul R. Bessell, Emily H. Crowley, Rian Snijders, Andrew Hope, Iñaki Tirados, Sophie Dunkley, Paul Verlé, Junior Lebuki, Chansy Shampa, Erick Mwamba Miaka, Fabrizio Tediosi, and Kat S. Rock. Cost-effectiveness of end-game strategies against sleeping sickness across the Democratic Republic of Congo. *medRxiv*, 2024.

[3] RE Crump, M Aliee, SA Sutherland, CI Huang, EH Crowley, SEF Spencer, MJ Keeling, C Shampa, Miaka E Mwamba, and KS Rock. Modelling timelines to elimination of sleeping sickness in the Democratic Republic of Congo, accounting for possible cryptic human and animal transmission. *Parasit Vectors*, 2024.

[4] Ronald E. Crump, Ching-I Huang, Edward S. Knock, Simon E. F. Spencer, Paul E. Brown, Erick Mwamba Miaka, Chansy Shampa, Matt J. Keeling, and Kat S. Rock. Quantifying epidemiological drivers of *gambiense* human African trypanosomiasis across the Democratic Republic of Congo. *PLOS Computational Biology*, 17(1):1–23, 01 2021.

[5] Christopher N. Davis, Ronald E. Crump, Samuel A. Sutherland, Simon E. F. Spencer, Alice Corbella, Shampa Chansy, Junior Lebuki, Erick Mwamba Miaka, and Kat S. Rock. Comparison of stochastic and deterministic models for *gambiense* sleeping sickness at different spatial scales: A health area analysis in the DRC. *PLOS Computational Biology*, 20(4):1–25, 04 2024.

[6] Rian Snijders, Alexandra P.M. Shaw, Richard Selby, Inaki Tirados, Paul R Bessell, Alain Fukinsia, Erick Miaka, Fabrizio Tediosi, Epco Hasker, and Marina Antillon. Extra words the cost of sleeping sickness vector control in the Democratic Republic of the Congo. *PLOS Neglected Tropical Diseases*, 2024.
